# Bacterial sexually transmitted infections and related antibiotic use among individuals eligible for doxycycline post-exposure prophylaxis in the United States

**DOI:** 10.1101/2025.02.24.25322788

**Authors:** Anna M. Parker, Jennifer J. Chang, Ligong Chen, Laura M. King, Sandra I. McCoy, Joseph A. Lewnard, Katia J. Bruxvoort

**Affiliations:** Division of Epidemiology, School of Public Health, University of California, Berkeley, Berkeley, California 94720, United States; Department of Infectious Diseases, Los Angeles Medical Center, Southern California Permanente Medical Group, Los Angeles, California, United States; Perisphere Real World Evidence, Austin, Texas 78751, United States; Gilead Sciences, Inc., Foster City, California 94404, United States; Division of Infectious Diseases & Vaccinology, School of Public Health, University of California, Berkeley, Berkeley, California 94720, United States; Center for Computational Biology, College of Computing, Data Science, and Society, University of California, Berkeley, Berkeley, California 94720, United States; Department of Epidemiology, University of Alabama at Birmingham, 1665 University Boulevard, Birmingham, AL, 35233, United States

**Keywords:** Doxycycline postexposure prophylaxis, doxyPEP, MSM, antibiotics

## Abstract

**Background:** Doxycycline postexposure prophylaxis (doxyPEP) can prevent bacterial sexually transmitted infections (STIs) among men who have sex with men (MSM) and transgender women. However, concern surrounds the volume of tetracycline use needed to realize these benefits, and whether potential risks of increased tetracycline exposure outweigh benefits of doxyPEP for specific populations.

**Methods:** We estimated incidence rates of gonorrhea, chlamydia, and syphilis and related antibiotic prescribing among commercially-insured US males and transgender individuals using the Merative MarketScan® Research Databases during 2016-2019. We evaluated potential impacts of doxyPEP implementation under risk-based prioritization schemes focusing on HIV pre-exposure prophylaxis (PrEP) recipients, people living with HIV (PLWH), and people with prior STI diagnoses.

**Results:** Incidence rates of gonorrhea, chlamydia, and syphilis among PLWH and PrEP recipients with ≥1 STI diagnosis in the prior year totaled 33.3-35.5 episodes per 100 person-years. Direct effects of doxyPEP could prevent 7.4-9.6 gonorrhea diagnoses, 7.3-8.1 chlamydia diagnoses, and 3.1-5.9 syphilis diagnoses per 100 person-years within these populations. Expected increases in tetracycline consumption resulting from doxyPEP implementation were equivalent to 271.9-312.9 additional 7-day doxycycline treatment courses (resembling current standards for chlamydia treatment) per 100 person-years of use. This increase corresponded to the equivalent of 36.5-37.0, 37.0-38.7, and 46.1-100.2 additional 7-day doxycycline treatment courses for each prevented chlamydia, gonorrhea, and syphilis episode, respectively. These increases in doxycycline use exceeded anticipated reductions in STI-related prescribing of cephalosporins, macrolides, and penicillins by 16-69 fold margins.

**Conclusions:** Estimates of changes in antibiotic use and STI incidence resulting from doxyPEP implementation in differing populations may inform priority-setting for this intervention.

## INTRODUCTION

Bacterial sexually transmitted infections (STIs) pose substantial burden due to their prevalence and potential sequelae. In the United Sates (US), >2.4 million gonorrhea, chlamydia, and syphilis cases were reported in 2023, and the number of reported syphilis cases was 80% higher than in 2018.^1^ Young people, men who have sex with men (MSM), transgender women (TGW), and people living with HIV (PLWH) are disproportionately affected by STIs.^2,3^

The US Centers for Disease Control and Prevention (CDC) recommended doxycycline post-exposure prophylaxis (doxyPEP) for prevention of bacterial STIs among certain populations in 2024 on the basis of shared clinical decision-making between patients and their healthcare providers.^4^ These recommendations encompass MSM and TGW who were diagnosed with a bacterial STI in the preceding 12 months. Doxycycline is typically well-tolerated and used for many acute bacterial infections.^5^ Longer-term courses are also prescribed for malaria prophylaxis and for skin conditions including moderate-to-severe inflammatory acne. DoxyPEP consists of 200 mg doxycycline taken orally within 72 hours (and ideally <24 hours) after sex.^6^ Recent trials have demonstrated that doxyPEP reduces chlamydia and syphilis infections among MSM, TGW, and PLWH by approximately 70%.^6–8^ Results were less promising for gonorrhea infections, with efficacy estimates spanning 17% in a French trial^6^ to 55% in a US trial.^7^

However, concerns surround possible contributions of doxyPEP to increased antibiotic use and antimicrobial resistance (AMR).^9^ In the US, approximately half of all *Neisseria gonorrhoeae* infections are resistant to ≥1 antibiotic;^10^ models suggest broad doxyPEP implementation may exacerbate this threat.^11^ Moreover, doxycycline exposure among carriers of *Staphylococcus aureus* and other commensal organisms has been reported to select for tetracycline resistance.^12–14^ While these considerations suggest doxyPEP implementation strategies should prioritize populations for whom the benefits of STI reduction outweigh potential risks associated with increased antibiotic consumption, evidence to inform such benefit-risk assessments is limited. Persons enrolled in randomized trials of doxyPEP,^15^ as well as observational studies recruiting patients in STI clinics,^16^ may not represent all populations eligible to receive doxyPEP under current recommendations. We analyzed real-world insurance claims data to assess the incidence of bacterial STIs and related antibiotic use that may be preventable through doxyPEP implementation. We estimated achievable reductions in STI incidence, and associated increases in doxycycline use, under differing doxyPEP implementation strategies.

## METHODS

### Study design, cohorts, and clinical outcomes

We conducted a retrospective cohort study using the Merative™ MarketScan® Research Databases, a US-wide collection of administrative and commercial healthcare data from adjudicated claims across multiple commercial insurance plans. The study population consisted of commercially-insured males and transgender individuals (**Table S1**) aged 18-64 years at any time between 1 January, 2017 and 31 December, 2019. We included time-at-risk for each individual following any 12-month period (extending as early as 1 January, 2016) during which they were continuously enrolled in both medical and prescription coverage (no lapse >60 days) and filled ≥1 prescription.

We used International Classification of Diseases, Tenth Revision (ICD-10) codes and National Drug Codes (NDCs) to identify individuals likely belonging population strata prioritized for doxyPEP implementation (**Tables S2**-**S6**). Our primary analyses encompassed persons receiving HIV pre-exposure prophylaxis (PrEP) and PLWH. To identify PrEP recipients, we used a previously-validated algorithm^17^ identifying individuals who filled prescriptions for tenofovir disoproxil and emtricitabine (TDF-FTC), excluding those with a needlestick exposure within ≤10 days of the prescription fill or any prior hepatitis B, HIV, or AIDS-defining opportunistic infection diagnoses. Whereas persons already receiving PrEP and PLWH may not encompass all individuals who could benefit from doxyPEP, we considered these populations as representing those most likely to adopt doxyPEP given their existing linkage to preventive sexual health services. Although MSM and non-MSM cannot be readily distinguished from administrative claims data, MSM comprise the vast majority of US males using PrEP^18^, and account for 87% of US males living with HIV.^19^

We also defined strata of individuals who had received ≥1, ≥2, or ≥3 diagnoses of chlamydia, gonorrhea, or syphilis in the preceding 12 months (as described below; **Tables S7**-**S9**). While both MSM and non-MSM could be represented among persons with prior bacterial STI diagnoses, MSM are over-represented within this population,^20^ and the clinical rationale for doxyPEP applies to all individuals at high risk of bacterial STIs, regardless of the gender of their sexual partners.

As PrEP use may be intermittent, we further sought to distinguish person-time at risk associated with periods during which individuals were expected to be using PrEP actively. Among all individuals who had received ≥1 PrEP fill, we distinguished follow-up occurring within 3 months after a new prescription fill as periods with “active” PrEP prescriptions, reflecting the typical 3-month supply per dispense. We also distinguished risk periods where individuals had received ≥3 PrEP fills within the preceding 12 months as periods where we expected individuals were engaged in “consistent” PrEP use.

Study outcomes were diagnoses and class-specific antibiotic fill-days for bacterial STIs (chlamydia, gonorrhea, and syphilis infections). We identified oral antibiotic prescriptions using NDCs with therapeutic classifications of 4 or 6-20, and injectable antibiotics via Healthcare Common Procedure Coding System (HCPCS) codes (**Table S10**). An antibiotic fill-day was defined as a day with ≥1 oral antibiotic fill or antibiotic injection. As syphilis treatment strategies frequently involve injections over multiple days, multiple fill-days could result from a single syphilis episode. We defined STI-associated fills with first- or second-line antibiotics (**Table S11**) as those occurring within 3 days before or after syphilis and gonorrhea diagnoses, and within 7 days before to 3 days after a chlamydia diagnosis (allowing a long period to capture antibiotics received prior to chlamydia diagnoses to account for presumptive treatment during gonorrhea co-infection). Chlamydia mono-infections were those occurring without accompanying gonorrhea infections within periods 3 days before to 7 days diagnosis.

### Descriptive analyses and baseline rates

We present descriptive characteristics of the study population as ranges of rates or proportions across cohorts eligible for analysis in 2016, 2017, and 2018. We calculated incidence rates (IRs) of chlamydia, gonorrhea, and syphilis, and IRs of antibiotic fill-days associated with these outcomes. We defined an aggregate IR for STI-related antibiotic fill-days associated with ≥1 syphilis, gonorrhea, or chlamydia diagnosis to avoid double-counting of overlapping treatment regimens targeting chlamydia/gonorrhea co-infection. We stratified rates for PrEP recipients, PLWH, and individuals with prior STI diagnoses, and computed subgroup-specific rates within each cohort according to individuals’ number of STI diagnoses in the preceding 12 months. We obtained IRs and accompanying 95% confidence intervals via generalized estimating equations with a Poisson link function.

### Modeling analyses

Within each population stratum, we estimated the potential direct effect of doxyPEP on the IR of each outcome. We parameterized direct effects of doxyPEP in preventing each infection, and first-line STI-related antibiotic fill-days for the same infection, using pathogen-specific relative-risk estimates from a recent meta-analysis of randomized controlled trials of doxyPEP.^21^ We did not consider treatment effects on IRs of STI-related antibiotic fill-days for second-line antibiotic classes; *N. gonorrhoeae* infections resistant to cephalosporins and second-line therapies are also typically resistant to tetracycline.^22^ As one doxyPEP trial^6^ did not report statistically-significant effects of doxyPEP on gonorrhea incidence, we conducted sensitivity analyses including effects on chlamydia and syphilis only.

We also projected possible increases in standardized tetracycline fill-days expected to result from doxyPEP use in each population stratum (**Text S1**). We estimated the rate of casual or first-time condomless anal sex partnerships among populations prioritized for doxyPEP implementation using data from a previous study of sexual encounter frequencies and characteristics among MSM. These data defined rates for MSM generally, and for MSM using PrEP and MSM with HIV.^23,24^ To account for expected differences in the frequency of sexual encounters across population strata with differing STI history, we used data from three studies^25–27^ measuring associations between MSM’s risk of bacterial STI diagnoses and their number of recent sex partners (**Table S12**; **Text S1**).

We harmonized comparisons of doxycycline consumption attributable to doxyPEP with estimated fill-days for acute STIs, equating each prophylactic use of 200mg doxycycline to one-seventh of an STI-associated tetracycline antibiotic fill-day. This standardization reflected the typical chlamydia treatment regimen of two 100mg doxycyline doses daily for 7 days. We quantified increases in consumption due to doxyPEP implementation by adding one standardized fill-day for every seven anticipated prophylactic uses of doxycycline after condomless anal sex with new or casual partners. We estimated incidence rate differences (IRDs) for tetracycline fill-days accounting for additional fill-days due to doxyPEP less STI-associated tetracycline fill-days prevented.

We also estimated potential population-level changes in STI incidence and antibiotic use assuming varying levels of doxyPEP uptake within each recipient stratum. We computed IRDs for diagnoses of gonorrhea, chlamydia, and syphilis as well as IRDs for fill-days for each antibiotic class, considering scenarios where doxyPEP is available for individuals with ≥1, ≥2, or ≥3 STI diagnoses in the prior year and where 30%, 50%, or 70% of eligible individuals adopt doxyPEP.

Last, to facilitate interpretation, we used our IRD estimates for STI diagnoses and antibiotic fill-days to quantify net changes in tetracycline consumption per STI diagnosis prevented by doxyPEP. This estimate can be interpreted as a “number needed to treat”^28^ for doxyPEP, defined as the net increase in standardized tetracycline fill-days per incident STI diagnosis prevented. We obtained this value by dividing IRDs for tetracycline fill-days by IRDs for gonorrhea, chlamydia, and syphilis diagnoses.

These analyses of deidentified insurance claims data were considered exempt from review by the University of Alabama, Birmingham Institutional Review Board and the Committee for the Protection of Human Subjects at the University of California, Berkeley.

## RESULTS

### Study population and baseline rates

Cohorts of PrEP recipients and PLWH increased from 10,679-15,384 individuals and from 8,095-11,011 individuals, respectively, between 2017-2019 (**Table S13**). Between 91.4-93.5% of PrEP recipients and PLWH had received no STI diagnoses in the preceding year; 1.1-1.9% had received ≥2 STI diagnoses. For alternate eligibility criteria based on STI history alone, we identified 5,621-7,609 and 825-1,249 individuals each year who, in the preceding year, had received ≥1 and ≥2 STI diagnoses, respectively.

Incidence rates of all three bacterial STIs totaled 10.8 and 8.1 diagnoses per 100 person-years among PrEP recipients and PLWH, respectively (**Table 1**). Restricting analysis cohorts to individuals who received ≥1 STI diagnosis in the preceding year, incidence rates of STI diagnoses totaled 33.3-35.5 per 100 person-years among these populations; rates of STI-related antibiotic fill-days per 100 person-years were 2.2-4.5 for cephalosporins, 1.6-3.5 for macrolides, 3.3-6.2 for penicillins, and 1.1-1.4 for tetracyclines (**Table 2**). These rates of STI-related antibiotic fill-days were roughly 3-fold higher among PrEP recipients and PLWH who received ≥1 STI diagnosis in the preceding year than among all PrEP recipients and PLWH. Overall rates of STI-related antibiotic use were highest among individuals aged 25-34 years, but similar across age groups within each risk stratum (**Tables S18**-**S21**). Most STI-related antibiotic fill-days were associated with gonorrhea infections (**Table S22-24**).

**Table 1:** Incidence rate of sexually transmitted infections among males and transgender individuals from 2017 to 2019 in the United States.

| Cohort |  | Incidence rate per 100-person years (95% CI) |  |  |  |
| --- | --- | --- | --- | --- | --- |
|  |  | *Any STI* | *Gonorrhea* | *Chlamydia* | *Syphilis* |
| PLWH<sup>1</sup> | All | 8.1 (7.7, 8.6) | 3.3 (3.1, 3.6) | 1.8 (1.7, 2.0) | 3.0 (2.8, 3.2) |
|  | ≥1 STI in year prior | 33.3 (30.0, 36.9) | 16.5 (14.5, 18.7) | 9.2 (7.6, 11.0) | 7.7 (6.6, 9.1) |
|  | ≥2 STIs in year prior | 61.2 (51.3, 72.8) | 32.0 (26.0, 39.3) | 18.7 (14.3, 24.6) | 10.5 (7.8, 14.0) |
|  | ≥3 STIs in year prior | 93.5 (71.3, 123.4) | 50.5 (37.2, 68.6) | 29.5 (18.5, 47.5) | 13.7 (8.3, 22.5) |
| PrEP use<sup>2</sup> | All | 10.8 (10.3, 11.2) | 5.9 (5.6, 6.2) | 3.1 (2.8, 3.3) | 1.8 (1.7, 2.0) |
|  | ≥1 STI in year prior | 35.5 (32.9, 38.5) | 21.4 (19.5, 23.5) | 10.0 (8.8, 11.5) | 4.1 (3.4, 4.8) |
|  | ≥2 STIs in year prior | 59.1 (51.7, 68.0) | 37.3 (31.9, 43.4) | 18.0 (14.6, 22.5) | 4.0 (2.8, 5.7) |
|  | ≥3 STIs in year prior | 82.6 (64.3, 106.2) | 49.8 (37.4, 66.2) | 29.2 (20.2, 42.2) | 4.0 (1.8, 8.6) |
| Active PrEP use<sup>3</sup> | All | 11.5 (11.0, 12.0) | 6.4 (6.1, 6.8) | 3.3 (3.0, 3.5) | 1.8 (1.7, 2.0) |
|  | ≥1 STI in year prior | 36.9 (34.1, 39.9) | 22.1 (20.2, 24.3) | 10.8 (9.6, 12.2) | 3.9 (3.3, 4.6) |
|  | ≥2 STIs in year prior | 59.6 (52.5, 68.0) | 36.3 (31.0, 42.5) | 18.4 (15.3, 22.3) | 4.9 (3.7, 6.5) |
|  | ≥3 STIs in year prior | 82.8 (65.5, 104.5) | 52.4 (39.7, 69.1) | 24.6 (17.7, 34.5) | 5.6 (3.3, 9.5) |
| Consistent PrEP use<sup>4</sup> | All | 13.0 (12.4, 13.6) | 7.3 (6.9, 7.7) | 3.7 (3.4, 4.0) | 2.1 (1.9, 2.3) |
|  | ≥1 STI in year prior | 43.3 (40.0, 46.9) | 25.6 (23.3, 28.3) | 13.7 (11.9, 15.6) | 4.0 (3.2, 4.8) |
|  | ≥2 STIs in year prior | 69.9 (61.2, 80.0) | 42.2 (36.1, 49.2) | 23.6 (19.0, 29.2) | 4.2 (2.8, 6.3) |
|  | ≥3 STIs in year prior | 107.9 (86.6, 133.9) | 64.6 (50.2, 83.0) | 39.8 (28.1, 55.8) | 3.2 (1.2, 8.8) |
| STI history | ≥1 STI in year prior | 17.8 (16.9, 18.6) | 8.5 (8.0, 9.1) | 7.1 (6.7, 7.7) | 2.1 (1.9, 2.3) |
|  | ≥2 STIs in year prior | 39.5 (36.2, 43.2) | 19.9 (17.9, 22.1) | 16.5 (14.6, 18.7) | 3.2 (2.7, 3.9) |
|  | ≥3 STIs in year prior | 82.3 (71.0, 95.9) | 39.9 (33.6, 47.6) | 36.7 (29.0, 46.3) | 5.9 (4.2, 8.4) |

CI: Confidence interval.
Incidence rates were calculated with an intercept only general equation model with a Poisson distribution. Yearly incidence rates are reported in **Tables S14-17**.
<sup>1</sup>People living with HIV (PLWH).
<sup>2</sup>PrEP users have filled ≥1 PrEP prescription in the previous year.
<sup>3</sup>Active PrEP users have ≥1 PrEP prescription in the previous three months.
<sup>4</sup>Consistent PrEP users have ≥1 PrEP prescription in the previous three months and ≥3 PrEP fills in the previous year.

**Table 2:** Incidence rate of oral and injection antibiotic fills related to sexually transmitted infections (STIs) among males and transgender individuals from 2017 to 2019 in the United States.

| Cohort |  | Incidence of antibiotic fill-days related to STIs per 100-person years (95% CI) |  |  |  |  |  |
| --- | --- | --- | --- | --- | --- | --- | --- |
|  |  | Cephalosporins | Macrolides | Penicillins | Tetracyclines | Quinolones | Aminoglycosides |
| PLWH<sup>1</sup> | All | 2.2 (2.0, 2.4) | 1.6 (1.4, 1.8) | 6.2 (5.7, 6.7) | 1.1 (0.9, 1.4) | 0 (0, 0.1) | 0 (0, 0) |
|  | ≥1 STI in year prior | 10.8 (9.3, 12.6) | 7.8 (6.1, 10.0) | 21.8 (18.9, 25.2) | 4.3 (2.9, 6.4) | 0.1 (0, 0.4) | 0 (0, 0) |
|  | ≥2 STIs in year prior | 19.3 (14.9, 24.8) | 12.6 (8.5, 19.1) | 23.1 (17.1, 31.2) | 7.1 (3.6, 14.7) | 0 (0, 0) | 0 (0, 0) |
|  | ≥3 STIs in year prior | 31.5 (21.7, 45.7) | 15.7 (8.9, 32.0) | 29.4 (16.8, 51.3) | 4.2 (1.1, 16.5) | 0 (0, 0) | 0 (0, 0) |
| PrEP use<sup>2</sup> | All | 4.5 (4.2, 4.8) | 3.5 (3.3, 3.8) | 3.3 (3.0, 3.7) | 1.4 (1.2, 1.6) | 0.1 (0, 0.1) | 0 (0, 0.1) |
|  | ≥1 STI in year prior | 15.9 (14.1, 18.0) | 11.1 (9.5, 13.1) | 9.4 (8.0, 11.1) | 4.1 (3.0, 5.7) | 0.1 (0.1, 0.3) | 0 (0, 0.2) |
|  | ≥2 STIs in year prior | 25.0 (19.7, 31.8) | 14.3 (10.7, 19.2) | 10.2 (7.5, 13.9) | 6.0 (3.3, 10.8) | 0.1 (0, 1) | 0 (0, 0) |
|  | ≥3 STIs in year prior | 35.7 (21.8, 58.4) | 16.5 (9.2, 30.2) | 10.6 (5.8, 19.3) | 9.2 (3.7, 23.4) | 0.7 (0.1, 4.6) | 0 (0, 0) |
| Active PrEP use<sup>3</sup> | All | 5.3 (5.0, 5.6) | 4.3 (3.9, 4.6) | 3.3 (3.0, 3.7) | 1.6 (1.3, 1.9) | 0.1 (0, 0.1) | 0 (0, 0.1) |
|  | ≥1 STI in year prior | 17.3 (15.2, 19.7) | 12.6 (10.7, 14.8) | 9.4 (8.0, 11.0) | 4.9 (3.5, 6.7) | 0.2 (0.1, 0.5) | 0 (0, 0.3) |
|  | ≥2 STIs in year prior | 25.3 (19.7, 32.5) | 16.4 (12.3, 22.0) | 10.4 (7.4, 14.6) | 6.9 (3.7, 12.8) | 0.3 (0.1, 1.3) | 0.2 (0, 1.2) |
|  | ≥3 STIs in year prior | 36.5 (21.6, 61.5) | 24.8 (15.2, 40.9) | 11.7 (6.5, 21.1) | 10.2 (4.0, 26.2) | 0 (0, 0) | 0 (0, 0) |
| Consistent PrEP use<sup>4</sup> | All | 5.5 (5.1, 5.9) | 4.5 (4.1, 4.9) | 3.6 (3.3, 4.1) | 1.7 (1.4, 2.0) | 0.1 (0.1, 0.1) | 0 (0, 0.1) |
|  | ≥1 STI in year prior | 17.7 (15.6, 20.0) | 13.3 (11.3, 15.8) | 9.4 (7.9, 11.2) | 5.0 (3.6, 7.2) | 0.2 (0.1, 0.5) | 0 (0, 0.3) |
|  | ≥2 STIs in year prior | 26.1 (20.5, 33.2) | 17.6 (13.1, 23.7) | 9.6 (6.6, 14.1) | 7.2 (3.7, 13.8) | 0.4 (0.1, 1.5) | 0.2 (0, 1.3) |
|  | ≥3 STIs in year prior | 39.8 (23.6, 67.0) | 27.1 (16.6, 44.6) | 10.3 (5.2, 20.5) | 9.5 (3.5, 26.5) | 0 (0, 0) | 0 (0, 0) |
| STI history | ≥1 STI in year prior | 5.5 (5.0, 5.9) | 4.9 (4.4, 5.4) | 6.0 (5.3, 6.7) | 2.1 (1.8, 2.5) | 0.1 (0, 0.1) | 0.1 (0, 0.2) |
|  | ≥2 STIs in year prior | 11.3 (9.7, 13.3) | 10.2 (8.4, 12.4) | 7.3 (6.1, 8.7) | 4.0 (2.9, 5.5) | 0.1 (0, 0.3) | 0 (0, 0.2) |
|  | ≥3 STIs in year prior | 24.2 (18.2, 32.2) | 16.9 (11.4, 25.5) | 11.6 (8, 16.7) | 8.1 (4.7, 14.2) | 0.2 (0, 1.4) | 0 (0, 0) |

CI: confidence interval.
<sup>1</sup>People living with HIV (PLWH).
<sup>2</sup>PrEP use: filled ≥1 PrEP prescription in the previous year.
<sup>3</sup>Active PrEP use: ≥1 PrEP prescription in the previous three months.
<sup>4</sup>Consistent PrEP use: ≥1 PrEP prescription in the previous three months and ≥3 PrEP fills in the previous year.

### Anticipated effects of doxyPEP

DoxyPEP was expected to prevent 2.7, 2.5, and 1.4 gonorrhea, chlamydia, and syphilis diagnoses, respectively, per 100 person-years of use among PrEP recipients (**Figure 1**). Within risk periods 0-3 months after a PrEP prescription fill or after a year of consistent PrEP use, doxyPEP would prevent 3.1-3.3 gonorrhea diagnoses, 2.9-3.0 chlamydia diagnoses, and 1.5-1.6 syphilis diagnoses per 100 person-years of use (**Table S25**). Considering only PrEP recipients who experienced ≥1 STI diagnosis in the prior year, avertible incidence totaled 9.6, 8.1, and 3.1 gonorrhea, chlamydia, and syphilis diagnoses, respectively, per 100 person-years of doxyPEP use. The avertible burden comprised 1.5-2.3 diagnoses of gonorrhea, chlamydia, and syphilis per 100 person-years within the general sample of PLWH, while among PLWH with ≥1 STI diagnosis in the preceding year, rates totaled 5.9-7.4 diagnoses per 100 person-years.

**Figure 1:**
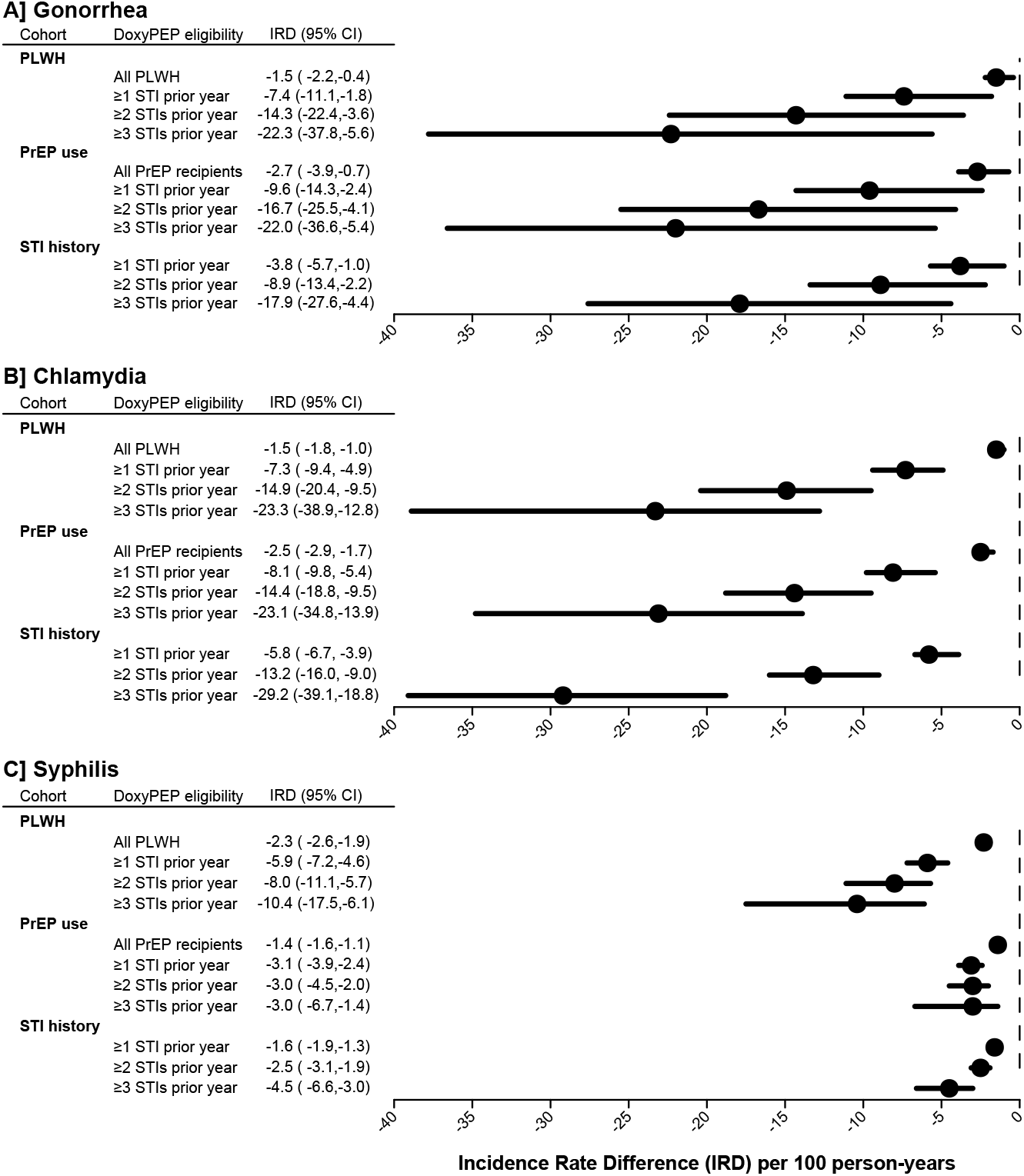
Incidence of gonorrhea, chlamydia, and syphilis preventable by direct effects of doxyPEP. We illustrate estimates of the incidence of gonorrhea (**A**), chlamydia (**B**), and syphilis (**C**) preventable by direct effects of doxyPEP among recipients within differing eligibility strata. Populations encompass males and transgender individuals living with HIV (PLWH), who receive HIV pre-exposure prophylaxis (PrEP), or with a history of STI diagnoses in the preceding year.

We estimated that PrEP recipients and PLWH would receive the equivalent of 203.5 and 141.0 additional tetracycline fill-days, respectively, per 100 person-years of doxyPEP use (**Table 3**), representing 128-145 fold increases over prevailing rates of STI-related tetracycline fill-days within these populations. Among PrEP recipients and PLWH with ≥1 STI diagnosis in the preceding year, corresponding increases were equivalent to 312.9 and 271.9 additional tetracycline fills, respectively, per 100 person-years of doxyPEP use, representing 63-76 fold increases over prevailing rates of STI-related tetracycline fills. Over 100 person-years of doxyPEP use, PrEP recipients and PLWH would experience 0.8-4.8 fewer STI-related fill-days or administrations for cephalosporins, macrolides, and penicillins, while PrEP recipients and PLWH who received ≥1 STI diagnosis in the prior year would experience 3.1-9.6 fewer STI-related fills of the same drugs.

**Table 3:** Incidence rate difference of oral and injection antibiotic fill days related to sexually transmitted infections among men and transgender individuals due to use of doxycycline post-exposure prophylaxis.

| Cohort |  | Incidence rate difference of antibiotic fills related to STIs per 100 person-years (95% CI) |  |  |  |
| --- | --- | --- | --- | --- | --- |
|  |  | *Cephalosporins* | *Macrolides* | *Penicillins* | *Tetracyclines* |
| PLWH<sup>1</sup> | All | -1.0 (-1.3, -0.3) | -0.8 (-1.0, -0.5) | -4.8 (-5.5, -3.8) | 141.0 (140.9, 141.2) |
|  | ≥1 STI in year prior | -4.9 (-6.1, -1.4) | -3.9 (-4.2, -2.5) | -16.7 (-20.2, -13.1) | 271.9 (270.9, 272.8) |
|  | ≥2 STIs in year prior | -8.7 (-9.9, -2.8) | -6.2 (-6.6, -4.8) | -17.7 (-24.4, -12.4) | 461.1 (457.3, 463.6) |
|  | ≥3 STIs in year prior | -14.2 (-14.9, -5.1) | -7.4 (-8.8, -5.9) | -22.4 (-39.6, -12.5) | 647.7 (638.1, 650.1) |
| PrEP use<sup>2</sup> | All | -2.0 (-2.8, -0.5) | -1.8 (-2.3, -1) | -2.6 (-0.3, -2.1) | 203.5 (203.4, 203.7) |
|  | ≥1 STI in year prior | -7.2 (-9.3, -2.0) | -5.8 (-6.6, -3.4) | -7.2 (-8.9, -5.6) | 312.9 (312.4, 313.5) |
|  | ≥2 STIs in year prior | -11.3 (-13, -3.6) | -7.6 (-8.1, -5.6) | -7.8 (-10.9, -5.5) | 421.4 (419.9, 422.8) |
|  | ≥3 STIs in year prior | -15.7 (-16.2, -6.6) | -8.9 (-11.1, -6.3) | -8.1 (-14.8, -4.3) | 532.4 (526.8, 535.4) |
| Active PrEP use<sup>3</sup> | All | -2.4 (-3.3, -0.6) | -2.2 (-2.7, -1.2) | -2.5 (-3.0, -2.0) | 203.4 (203.3, 203.6) |
|  | ≥1 STI in year prior | -7.8 (-10.1, -2.2) | -6.7 (-7.5, -4.1) | -7.2 (-8.8, -5.6) | 312.4 (311.8, 313.2) |
|  | ≥2 STIs in year prior | -11.4 (-13.0, -3.7) | -8.6 (-9.2, -6.2) | -7.9 (-11.4, -5.4) | 420.7 (418.5, 422.5) |
|  | ≥3 STIs in year prior | -16.0 (-16.5, -6.9) | -12.7 (-13.9, -10.0) | -8.9 (-16.2, -4.8) | 531.5 (524.8, 535.1) |
| Consistent PrEP users<sup>4</sup> | All | -2.5 (-3.4, -0.7) | -2.3 (-2.8, -1.2) | -2.8 (-3.3, -2.2) | 203.4 (203.2, 203.5) |
|  | ≥1 STI in year prior | -8.0 (-10.3, -2.2) | -7.0 (-7.9, -4.2) | -7.2 (-8.9, -5.6) | 312.3 (311.6, 313.1) |
|  | ≥2 STIs in year prior | -11.8 (-13.5, -3.7) | -9.1 (-9.7, -6.5) | -7.4 (-10.9, -4.8) | 420.6 (418.2, 422.5) |
|  | ≥3 STIs in year prior | -17.4 (-18.0, -7.5) | -13.8 (-15.2, -10.9) | -7.9 (-15.7, -3.9) | 532.2 (525.0, 535.5) |
| STI history | ≥1 STI in year prior | -2.5 (-3.3, -0.7) | -2.7 (-3.1, -1.7) | -4.6 (-5.4, -3.6) | 174.6 (174.5, 174.9) |
|  | ≥2 STIs in year prior | -5.1 (-6.4, -1.5) | -5.7 (-6.2, -4.1) | -5.6 (-6.9, -4.3) | 299.2 (298.7, 299.7) |
|  | ≥3 STIs in year prior | -10.9 (-12.1, -3.6) | -9.4 (-10.7, -7.6) | -8.9 (-13.0, -5.9) | 447.2 (444.6, 449.2) |

CI: Confidence interval.
<sup>1</sup>People living with HIV (PLWH).
<sup>2</sup>PrEP use: filled ≥1 PrEP prescription in the previous year.
<sup>3</sup>Active PrEP use: ≥1 PrEP prescription in the previous three months.
<sup>4</sup>Consistent PrEP use: ≥1 PrEP prescription in the previous three months and ≥3 PrEP fills in the previous year.

We obtained similar results using differing estimates of the frequency with which individuals were expected to take doxyPEP based on modeled relationships between STI history and sexual risk (**Table S26**). Reductions in STI incidence were more modest when assuming no effect of doxyPEP on gonorrhea (**Table S27**).

### Number-needed-to-treat for STI prevention via doxyPEP

DoxyPEP implementation would result in the equivalent of 76.0-93.9, 82.5-95.7, and 61.0-145.3 additional STI-related tetracycline fill-days for each prevented episode of gonorrhea, chlamydia, and syphilis, respectively, among all PrEP recipients and PLWH (**Figure 2**; **Table S28**). For individuals within these strata who received ≥1 STI diagnosis in the preceding year, one case of gonorrhea, chlamydia, and syphilis could be prevented with the equivalent of 32.2-36.5, 37.0-38.7, and 46.1-100.2 additional tetracycline fill-days, respectively. Increases in fill-days needed to prevent each gonorrhea and chlamydia episode were only modestly lower among PrEP recipients and PLWH who received ≥2 or ≥3 STI diagnoses in the prior year.

**Figure 2:**
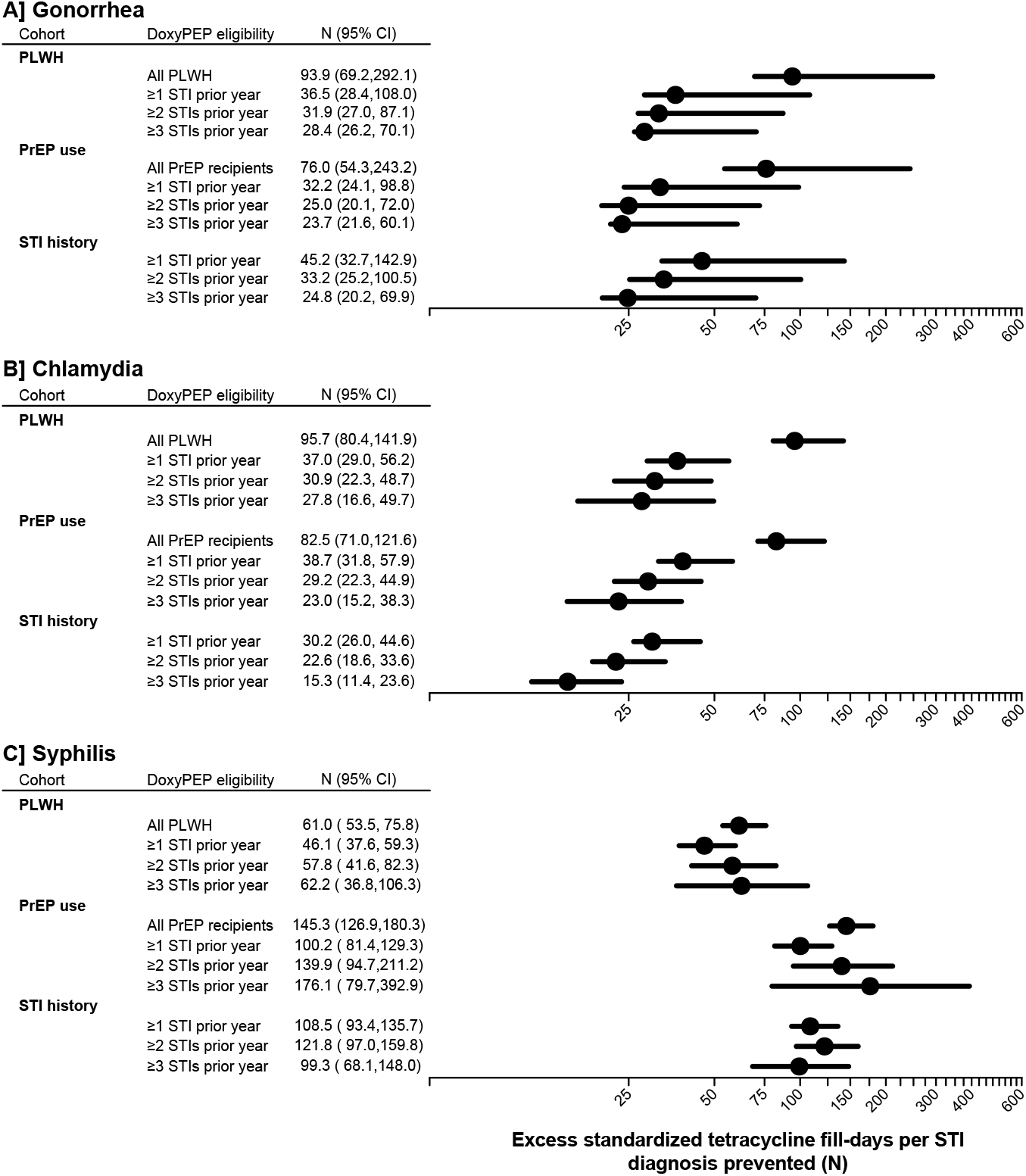
Number-needed-to-treat for STI prevention via doxyPEP. We illustrate estimates of the excess tetracycline fill-days (standardized to resemble a 7-day treatment course for acute STI management) expected to occur for each case of gonorrhea (**A**), chlamydia (**B**), and syphilis (**C**) prevented by doxyPEP. Populations encompass males and transgender individuals living with HIV (PLWH), who receive HIV pre-exposure prophylaxis (PrEP), or with a history of STI diagnoses in the preceding year.

Increases in tetracycline consumption associated with doxyPEP use among all PrEP recipients or PLWH corresponded to the equivalent of 100.1-140.8, 111.1-171.7, and 29.7-80.2 additional tetracycline fill-days per averted STI-related course of treatment with cephalosporins, macrolides, and penicillins, respectively (**Table 4**). Restricting doxyPEP to PrEP recipients and PLWH with ≥1 STI diagnosis in the prior year, we estimated the equivalent of 43.4-55.5, 54.1-69.0, and 16.2-43.3 additional tetracycline fill-days per averted STI-related course of treatment with the same drugs.

**Table 4:**
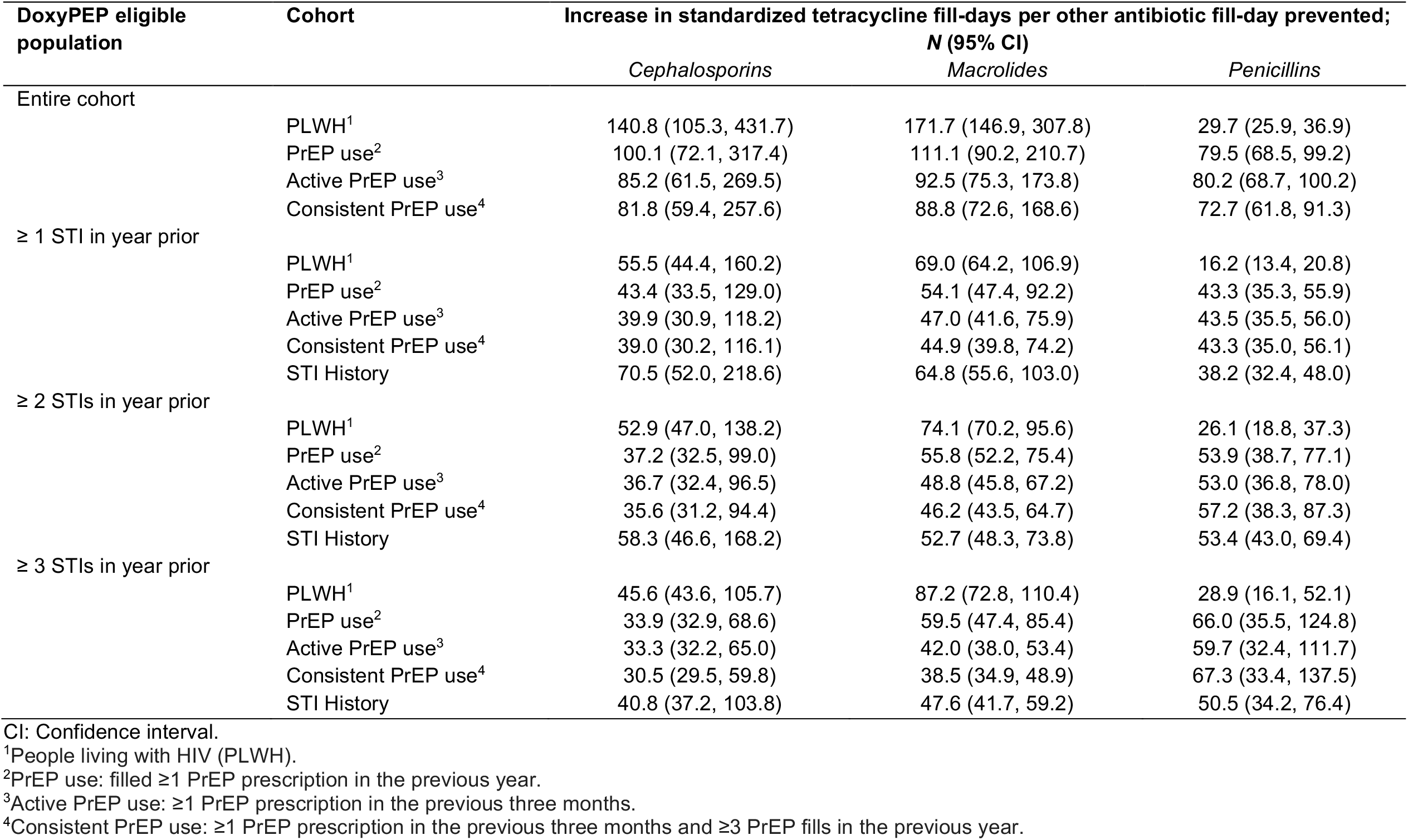
Estimated increase in standardized tetracycline fills per STI-related antibiotic prevented by use of doxycycline post-exposure prophylaxis.

### Overall impact of implementation

With 50% uptake among PrEP recipients, doxyPEP would prevent 22.0%, 38.7%, and 22.6% of all diagnoses of gonorrhea, chlamydia, and syphilis, respectively, among PrEP recipients (**Table 5**). Use among 50% of PrEP recipients with ≥1 prior STI would prevent 0.4, 0.3, and 0.1 diagnoses, representing only 5.1-6.8% of diagnoses with these conditions among all PrEP recipients. Uptake among 50% of all PrEP recipients or 50% of PrEP recipients with ≥1 STI diagnosis in the prior year would increase tetracycline use by the equivalent of 101.8 and 12.7 fills per 100 person-years, respectively, representing 73-fold and 9.1-fold increases over prevailing rates of STI-related tetracycline fills among all PrEP recipients. Restricting doxyPEP to individuals with ≥2 or ≥3 STI diagnoses in the prior year would prevent <6% of diagnoses for each STI among all PrEP recipients, while increasing STI-related tetracycline fill-days by the equivalent of 264% and 71% all PrEP recipients.

**Table 5:** Estimated overall cohort incidence rate difference of sexually transmitted infections and standardized tetracycline fill-days among males and transgender individuals by doxycycline postexposure prophylaxis uptake and implementation strategy.

| Uptake scenario | Cohort | DoxyPEP-eligible population | Incidence rate difference per 100 person-years (95% CI) |  |  |  |
| --- | --- | --- | --- | --- | --- | --- |
|  |  |  | Gonorrhea | Chlamydia | Syphilis | Tetracycline fill-days |
| 50% uptake | PLWH | All | -0.7 (-1.1, -0.2) | -0.7 (-0.9, -0.5) | -1.2 (-1.3, -0.9) | 70.5 (70.5, 70.6) |
|  |  | ≥1 STI in year prior | -0.2 (-0.3, -0.1) | -0.2 (-0.3, -0.2) | -0.2 (-0.2, -0.2) | 9.0 (9.0, 9.0) |
|  |  | ≥2 STIs in year prior | -0.1 (-0.1, 0) | -0.1 (-0.1, -0.1) | 0 (-0.1, 0) | 2.9 (2.8, 2.9) |
|  |  | ≥3 STIs in year prior | 0 (0, 0) | 0 (-0.1, 0) | 0 (0, 0) | 1.1 (1.0, 1.1) |
|  | PrEP user | All | -1.3 (-2.0, -0.3) | -1.2 (-1.4, -0.8) | -0.7 (-0.8, -0.6) | 101.8 (101.7, 101.8) |
|  |  | ≥1 STI in year prior | -0.4 (-0.5, -0.1) | -0.3 (-0.4, -0.2) | -0.1 (-0.2, -0.1) | 12.7 (12.7, 12.8) |
|  |  | ≥2 STIs in year prior | -0.1 (-0.2, 0) | -0.1 (-0.2, -0.1) | 0 (0, 0) | 3.7 (3.7, 3.8) |
|  |  | ≥3 STIs in year prior | 0 (0, 0) | 0 (-0.1, 0) | 0 (0, 0) | 1.1 (1.0, 1.1) |
|  | STI history | ≥1 STI in year prior | -1.9 (-2.8, -0.5) | -2.9 (-3.3, -2.0) | -0.8 (-0.9, -0.6) | 87.3 (87.2, 87.4) |
|  |  | ≥2 STIs in year prior | -0.7 (-0.9, -0.2) | -1 (-1.3, -0.7) | -0.2 (-0.2, -0.1) | 23.4 (23.4, 23.4) |
|  |  | ≥3 STIs in year prior | -0.2 (-0.3, -0.1) | -0.4 (-0.5, -0.2) | -0.1 (-0.1, 0) | 5.5 (5.5, 5.5) |
| 30% uptake | PLWH | All | -0.4 (-0.7, -0.1) | -0.4 (-0.5, -0.3) | -0.7 (-0.8, -0.6) | 42.3 (42.3, 42.4) |
|  |  | ≥1 STI in year prior | -0.1 (-0.2, 0.0) | -0.1 (-0.2, -0.1) | -0.1 (-0.1, -0.1) | 5.4 (5.4, 5.4) |
|  |  | ≥2 STIs in year prior | 0.0 (0.0, 0.0) | 0.0 (-0.1, 0.0) | 0.0 (0.0, 0.0) | 1.7 (1.7, 1.7) |
|  |  | ≥3 STIs in year prior | 0.0 (0.0, 0.0) | 0.0 (0.0, 0.0) | 0.0 (0.0, 0.0) | 0.6 (0.6, 0.6) |
|  | PrEP user | All | -0.8 (-1.2, -0.2) | -0.7 (-0.9, -0.5) | -0.4 (-0.5, -0.3) | 61.1 (61.0, 61.1) |
|  |  | ≥1 STI in year prior | -0.3 (-0.3, -0.1) | -0.2 (-0.3, -0.1) | -0.1 (-0.1, 0.0) | 7.6 (7.6, 7.7) |
|  |  | ≥2 STIs in year prior | -0.1 (-0.1, 0.0) | -0.1 (-0.1, 0.0) | 0.0 (0.0, 0.0) | 2.2 (2.2, 2.2) |
|  |  | ≥3 STIs in year prior | 0.0 (0.0, 0.0) | 0.0 (0.0, 0.0) | 0.0 (0.0, 0.0) | 0.6 (0.6, 0.6) |
|  | STI history | ≥1 STI in year prior | -1.2 (-1.7, -0.3) | -1.7 (-2, -1.2) | -0.5 (-0.6, -0.4) | 52.4 (52.3, 52.5) |
|  |  | ≥2 STIs in year prior | -0.4 (-0.6, -0.1) | -0.6 (-0.8, -0.4) | -0.1 (-0.1, -0.1) | 14.1 (14.0, 14.1) |
|  |  | ≥3 STIs in year prior | -0.1 (-0.2, 0.0) | -0.2 (-0.3, -0.1) | 0.0 (0.0, 0.0) | 3.3 (3.3, 3.3) |
| 70% uptake | PLWH | All | -1.0 (-1.5, -0.3) | -1.0 (-1.2, -0.7) | -1.6 (-1.8, -1.3) | 98.7 (98.6, 98.8) |
|  |  | ≥1 STI in year prior | -0.3 (-0.4, -0.1) | -0.3 (-0.4, -0.2) | -0.3 (-0.3, -0.2) | 12.6 (12.5, 12.6) |
|  |  | ≥2 STIs in year prior | -0.1 (-0.1, 0) | -0.1 (-0.2, -0.1) | -0.1 (-0.1, 0) | 4.0 (4.0, 4.0) |
|  |  | ≥3 STIs in year prior | -0.1 (-0.1, 0) | -0.1 (-0.1, 0) | 0 (0, 0) | 1.5 (1.5, 1.5) |
|  | PrEP user | All | -1.9 (-2.7, -0.5) | -1.7 (-2, -1.2) | -1.0 (-1.1, -0.8) | 142.5 (142.4, 142.6) |
|  |  | ≥1 STI in year prior | -0.6 (-0.7, -0.2) | -0.5 (-0.6, -0.3) | -0.2 (-0.2, -0.1) | 17.8 (17.8, 17.9) |
|  |  | ≥2 STIs in year prior | -0.2 (-0.3, -0.1) | -0.2 (-0.2, -0.1) | 0 (-0.1, 0) | 5.2 (5.2, 5.2) |
|  |  | ≥3 STIs in year prior | -0.1 (-0.1, 0) | -0.1 (-0.1, 0) | 0 (0, 0) | 1.5 (1.4, 1.5) |
|  | STI history | ≥1 STI in year prior | -2.7 (-4.0, -0.7) | -4.0 (-4.7, -2.7) | -1.1 (-1.3, -0.9) | 122.2 (122.1, 122.4) |
|  |  | ≥2 STIs in year prior | -1.0 (-1.3, -0.3) | -1.5 (-1.8, -1) | -0.3 (-0.3, -0.2) | 32.8 (32.7, 32.8) |
|  |  | ≥3 STIs in year prior | -0.3 (-0.4, -0.1) | -0.5 (-0.7, -0.3) | -0.1 (-0.1, -0.1) | 7.7 (7.7, 7.8) |

CI: Confidence interval.
Incidence rate difference estimates can be interpreted as the cohort-wide change in STI-related antibiotic fills within each cohort for each uptake scenario, with doxyPEP targeted to the indicated eligible population. The STI history cohort contains all individuals with at least one STI in the prior year.
<sup>1</sup>People living with HIV (PLWH).
<sup>2</sup>PrEP use: filled ≥1 PrEP prescription in the previous year.
<sup>3</sup>Active PrEP use: ≥1 PrEP prescription in the previous three months.
<sup>4</sup>Consistent PrEP use: ≥1 PrEP prescription in the previous three months and ≥3 PrEP fills in the previous year.

Similarly, 50% uptake of doxyPEP among all PLWH would prevent 0.7, 0.7, and 1.2 gonorrhea, chlamydia, and syphilis diagnoses, respectively, per 100 person-years; 50% uptake among PLWH with ≥1 STI diagnosis in the prior year would prevent 0.2 diagnoses of each STI per 100 person-years, representing 6.1-11.1% of such diagnoses among all PLWH. These scenarios would increase tetracycline use among PLWH by the equivalent of 70.5 and 9.0 fill-days per 100 person-years.

## DISCUSSION

Using doxyPEP to prevent STIs among commercially-insured individuals currently eligible to receive this intervention would require the equivalent of 32.2-36.5, 37.0-38.7, and 46.1-100.2 additional standardized 7-day tetracycline courses for each prevented episode of gonorrhea, chlamydia, and syphilis, respectively. These increases in tetracycline consumption surpasses decreases in STI-related consumption of other antibiotics by factors of 43.4-100.2 within the prioritized populations.

Whereas current guidelines recommend doxyPEP for individuals with STI diagnosis in the past year, early adopters of doxyPEP in several recent real-world studies have also included individuals without recent STI history. In one US study, 63% of doxyPEP recipients had not been diagnosed with any STI in the preceding year and 15% had never been diagnosed with an STI;^29^ in a second, 52% of doxyPEP adopters had no STI diagnoses in the preceding year.^30^ In studies in Italy^31^ and Spain,^32^ 41-51%, 45-62% and 76-86% of doxyPEP recipients had never been diagnosed with gonorrhea, chlamydia, and syphilis, respectively. Prevailing doxyPEP use patterns may thus encompass populations with low baseline STI incidence.

Restricting doxyPEP to narrower risk groups, such as those with ≥2 or ≥3 STI diagnoses in the preceding year, only modestly lowered the estimated number of tetracycline fill-days needed to prevent one diagnosis of gonorrhea or chlamydia. Even with only 50% uptake among persons with ≥2 or ≥3 STI diagnoses in the preceding year, doxyPEP would increase tetracycline consumption by 71-264% among all PrEP recipients and PLWH in the studied populations. These limited gains in efficiency reflect the fact that individuals’ recent STI history is an imperfect predictor of future risk.

Commercially-insured PrEP recipients and PLWH within our cohort likely represent the populations who will receive doxyPEP under real-world conditions of implementation. In recent studies, 58-100% of doxyPEP adopters were already using PrEP or living with HIV.^29–31^ However, one US study reported that 9% of males in a cohort of early doxyPEP adopters identified as heterosexual, and 14% reported no history of anal sex with male partners in the preceding 12 months.^30^ Within our analyses, PrEP recipients and PLWH accounted for only 31.2-34.0% of all males who received ≥2 STI diagnoses in any preceding 1-year period. These findings identify a population at high risk of STIs who are not linked to sexual health services through PrEP or HIV care, among whom the clinical rationale for doxyPEP resembles that among populations currently recommended to receive doxyPEP.

In a previous modeling study using data from MSM and TGW who received care from a center specializing in sexual health services, doxyPEP implementation was expected to prevent 25.6-28.5 STI diagnoses per 100 person-years among PrEP recipients and PLWH.^16^ Our lower estimates of preventable burden correspond to the substantially lower rates of STI diagnoses observed among PrEP recipients and PLWH in a nationwide sample of commercially-insured individuals in comparison to STI clinic patients. Secondary analyses based on the same STI clinic-based cohort have suggested that even within this study population, increases in tetracycline consumption due to doxyPEP implementation exceeded reductions in antibiotic treatment of STIs by a factor of 42.^15^

Our study has several limitations. First, we do not consider population-level (indirect) effects of doxyPEP in reducing STI transmission. However, a prior modeling study estimated that such benefits could be offset by increases in tetracycline resistance, with doxyPEP expected to lose clinical effectiveness against gonorrhea within 2-12 years.^33^ Decreases in population-level STI transmission due to doxyPEP would reduce the individual-level clinical benefit that our study estimates is achievable through doxyPEP use, increasing the number of doxyPEP courses needed to prevent one STI episode. Second, claims data offer limited sensitivity in representing sexual and gender-minority populations, and do not enable restriction to MSM and TGW. However, real-world use of doxyPEP has not been strictly limited to MSM and TGW.^29^ Third, antibiotic resistance profiles vary geographically and may influence both prescribing practices and doxyPEP effectiveness. Fourth, our analyses may not fully capture STI-related antibiotic dispenses if some antibiotics are purchased without insurance reimbursement, received via expedited partner therapy, or obtained through informal sources. Fifth, we assumed one doxycycline dose for every casual or first-time condomless anal sex partnership. However, as doxyPEP is also recommended after condomless vaginal or oral sexual exposures, estimates may reflect lower bounds for real-world use. Finally, our cohort was limited to individuals enrolled in commercial insurance plans. Prior studies have demonstrated that lack of insurance is an important predictor of non-adoption or discontinuation of PrEP among US MSM,^34–36^ suggesting our sample may represent the populations most likely to receive doxyPEP under real-world conditions. High STI risk and lack of insurance among disadvantaged groups remain important health equity considerations,^37^ with doxyPEP awareness currently low among rural MSM and those without postsecondary education.^38^

Our findings offer a quantitative basis for evaluating potential benefit-risk tradeoffs of differing doxyPEP implementation strategies. Benefits of doxyPEP alongside STI prevention may include alleviation of the medical costs, productivity losses, and stigma associated with STIs. However, preliminary data have identified increased prevalence of tetracycline resistance in *S. aureus* and the gut microbiome^14^ among doxyPEP recipients,^7^ and in *S. aureus* infections within populations eligible for doxyPEP use.^12^ Ongoing surveillance for antimicrobial resistance in *N. gonorrhoeae* and other bacterial pathogens^39^ remains a priority to identify changes with implications for clinical care.

## Supporting information

Text S1, Tables S1-S28, Supplemental references

## Data Availability

This study used data from the Merative MarketScan Research Databases. Data use agreements between the authors' institutions and the data providers do not allow for secondary data sharing by the auathors.

