## Supplementary material for "Bacterial sexually transmitted infections and related antibiotic use among individuals eligible for doxycycline post-exposure prophylaxis in the United States": Text S1, Tables S1-S28, Supplemental references

**Contents of this supplement**

| Item | Title | Page |
|---|---|---|
| Text S1 | Supplemental methods. | 1 |
| Table S1 | Codes used to identify transgender persons. | 5 |
| Table S2 | National Drug Codes used to identify Emtricitabine/Tenofovir use. | 6 |
| Table S3 | Codes used to identify HIV infection. | 7 |
| Table S4 | Codes used to identify HIV-associated opportunistic infections. | 8 |
| Table S5 | Codes used to identify Hepatitis B infection. | 9 |
| Table S6 | Codes used to identify needlestick exposure. | 10 |
| Table S7 | Codes used to identify gonococcal infection. | 11 |
| Table S8 | Codes used to identify chlamydial infection. | 12 |
| Table S9 | Codes used to identify syphilis infection. | 13 |
| Table S10 | Healthcare Common Procedure Coding System (HCPCS) codes used to identify injectable antibiotics related to sexually transmitted infections. | 14 |
| Table S11 | Oral and injection antibiotics with first- or second-line classification used to treat sexually transmitted infections in the United States 2017 to 2019. | 15 |
| Table S12 | Estimated annual rates of new or casual unprotected anal sex partnerships among MSM according to risk characteristics. | 16 |
| Table S13 | Demographic characteristics of the study population, by risk stratum. | 17 |
| Table S14 | Incidence rate of sexually transmitted infections among males and transgender individuals stratified by year and over the entire study period. | 18 |
| Table S15 | Incidence rate of gonorrhea diagnoses among males and transgender individuals stratified by year. | 19 |
| Table S16 | Incidence rate of chlamydia diagnoses among males and transgender individuals stratified by year. | 20 |
| Table S17 | Incidence rate of syphilis diagnoses among males and transgender individuals stratified by year. | 21 |

| | | |
|---|---|---|
| Table S18 | Incidence rate of oral and injection antibiotic fills related to sexually transmitted infections (STIs) among males and transgender individuals aged 18 to 24 years from 2017 to 2019 in the United States. | 22 |
| Table S19 | Incidence rate of oral and injection antibiotic fills related to sexually transmitted infections (STIs) among males and transgender individuals aged 25 to 34 years from 2017 to 2019 in the United States. | 23 |
| Table S20 | Incidence rate of oral and injection antibiotic fills related to sexually transmitted infections (STIs) among males and transgender individuals aged 35 to 44 years from 2017 to 2019 in the United States. | 24 |
| Table S21 | Incidence rate of oral and injection antibiotic fills related to sexually transmitted infections (STIs) among males and transgender individuals aged 45 to 65 years from 2017 to 2019 in the United States. | 25 |
| Table S22 | Incidence rate of oral and injection antibiotic fills related to gonorrhea diagnoses among males and transgender individuals from 2017 to 2019 in the United States. | 26 |
| Table S23 | Incidence rate of oral and injection antibiotic fills related to chlamydia diagnoses with and without a gonorrhea co-infection among males and transgender individuals from 2017 to 2019 in the United States. | 27 |
| Table S24 | Incidence rate of oral and injection antibiotic fills related to syphilis among males and transgender individuals diagnoses from 2017 to 2019 in the United States. | 28 |
| Table S25 | Incidence rate difference of sexually transmitted infections among males and transgender individuals with 100% doxycycline postexposure prophylaxis (doxyPEP) uptake. | 29 |
| Table S26 | Estimates of incidence rate difference in standardized tetracycline fills with 100% doxycycline postexposure prophylaxis (doxyPEP) uptake utilizing alternative estimates of sexual encounter frequencies. | 30 |
| Table S27 | Incidence rate difference of oral and injection antibiotic fills related to sexually transmitted infections among males and transgender individuals with doxycycline postexposure prophylaxis having no effect on gonorrhea infections. | 31 |
| Table S28 | Estimated increase in standardized tetracycline fills per sexually transmitted infection prevented. | 32 |
| Supplemental references | | 33 |

**Text S1: Supplemental methods.**

To estimate the frequency with which MSM in differing risk strata may use doxyPEP, we used data from previous studies quantifying rates of unprotected anal sex partnerships with new or casual partners among MSM and studies measuring associations between MSM's recent numbers of such partnerships and their risk of bacterial STI diagnoses.

*Reference population*

We considered the sample enrolled in the ARTnet study as a reference population for quantifying rates of sexual partnership formation.[1,2] The sample for this study was recruited online among MSM between July 2017 and January 2019 directly after participation in the American Men's Internet Study,[3] an annual online behavioral survey recruiting MSM via email blasts and banner advertisements on websites or mobile phone applications targeting differing audiences (gay social networking, gay general interest, general social networking, and geospatial social networking). Consistency of sexually-transmitted infection incidence within this study sample and US surveillance data suggest risk characteristics of the population recruited in the ARTnet are representative of those of MSM in general. Specifically, self-reported HIV prevalence within the study sample was 10.8%, comparable to the estimate of 11.1% prevalence of diagnosed HIV infection among all US MSM.[4] Gonorrhea incidence within the sample was 5.07 cases/100 person-years, comparable to the estimate of 5.17 cases/100 person-years among MSM.[5] Additionally, prior comparative studies have reported that sexual risk behaviors are similar in MSM samples recruited through such online surveys and those recruited via physical venue-based sampling [6].

To account for the right-tailed shape of the distribution of individuals' annual reported partnerships, we considered this count to follow a negative binomial distribution, $Z_j \sim \text{Negative Binomial}(p_j, r_j)$ within strata ($j$) comprised of MSM using PrEP, MSM with HIV, and the general MSM population. We fit parameters $p_j$ and $r_j$ via least-squares fitting against reported distribution quantiles for each stratum within ARTnet study publications.[1,2] We sampled from the fitted distributions of the number of partnerships among pseudo-populations of 1 million individuals for each stratum to generate a random variable of individuals' annual partnership counts, $Z(i)$.

*Partnership counts by STI history*

We identified three US-based studies reporting data on STI risk and numbers of partnerships over differing recall periods among MSM.[7–9] The cohort study we used to parameterize our primary analysis model[8] reported annual incidence rates of gonorrhea and chlamydia urethral or rectal infections across strata of individuals with 1, 2-5, 6-10, 11-50, or ≥50 partners over a 6-month observation period. Using $Z(i)/2$ to measure individuals' expected number of partnerships over a 6-month period within the pseudo-population, we generated individual-specific rate parameters for STI diagnoses $\lambda(i)$ corresponding to their 6-month history of sexual partnerships:

$$\lambda(i) = \begin{cases} 0.0658 & \text{if } Z(i)/2 < 2 \\ 0.1192 & \text{if } 2 \leq Z(i)/2 < 6 \\ 0.1918 & \text{if } 6 \leq Z(i)/2 < 10 \\ 0.244 & \text{if } 10 \leq Z(i)/2 < 50 \\ 0.3385 & \text{if } Z(i)/2 \geq 50 \end{cases}$$

In turn, we sampled the number of STI diagnoses among individuals over a given year for our pseudo-population as

$$Y(i) \sim \text{Pois}(\lambda(i)).$$

We used the resulting draws of $Y$ to obtain conditional distributions of individuals' annual number of partnerships, $Z$, for strata with ≥1, ≥2, and ≥3 STI diagnoses in the prior year ($Y \geq 1$, $Y \geq 2$, and $Y \geq 3$; **Table S12**).

A second study reported numbers of partnerships in the preceding 12 months among MSM testing positive for gonorrhea or chlamydia to those testing negative in a US national sample.[7] To recover distributions of encounter rates among MSM expected to experience ≥1, ≥2, or ≥3 bacterial STIs annually, we first parameterized the relative risk of STI diagnosis (assumed to be equivalent to the incidence rate ratio of STI diagnoses) per additional sexual encounter. We fit Gamma distributions to reported 50%, 25%, and 75% quantiles of individuals' partnerships per year among those who tested positive for gonorrhea or chlamydia and those testing negative for both to recover rates of sexual partnership formation:

$$Z^*_{\text{STI}=1} \sim \text{Gamma}(\alpha_1, \beta_1)$$

$$Z^*_{\mathrm{STI}=0} \sim \mathrm{Gamma}(\alpha_0, \beta_0)$$

We sampled one million draws from the distribution of each rate, and used these draws from the resulting pseudo-population of individuals with or without STI diagnoses to estimate the relative risk via Poisson regression. We used the resulting regression coefficients to project individual-specific rates of STI diagnosis, $\lambda(i)$, as a function of their rate of partnership formation for the full pseudo-population,

$$\lambda(i) = \exp[\beta_0 + \beta_1 Z(i)].$$

In turn, we defined individuals' number of STI diagnoses in a given year, $Y(i)$, as a Poisson random variable parameterized by the individual-specific rate $\lambda(i)$,

$$Y(i) \sim \mathrm{Pois}\big(\lambda(i)\big).$$

Finally, a third cohort study[9] presented a continuous relationship of cumulative risk of a gonorrhea or chlamydia diagnosis over a 6-month period with respect to their number of casual partners over the same period. We digitized figures presenting this relationship to define the joint distribution of $\lambda/2$ with respect to $Z/2$, and sampled individuals' number of STI diagnoses in a given 12-month period as

$$Y(i) \sim \mathrm{Pois}\big(\lambda(i)\big).$$

**Table S1: Codes used to identify transgender persons.**

| Code type | Code | Code description |
|---|---|---|
| ICD10 | F64 | Gender identity disorders |
| ICD10 | F64.0 | Transsexualism |
| ICD10 | F64.1 | Gender identity disorder in adolescence and adulthood |
| ICD10 | F64.2 | Gender identity disorder in childhood |
| ICD10 | F64.8 | Other gender identity disorders |
| ICD10 | F64.9 | Gender identity disorder, unspecified |
| ICD10 | Z87.890 | Personal history of sex reassignment |
| ICD10-PCS | 0W4M070 | Creation of Vagina in Male Perineum with Autologous Tissue Substitute, Open Approach. |
| ICD10-PCS | 0W4M0J0 | Creation of Vagina in Male Perineum with Synthetic Substitute, Open Approach |
| ICD10-PCS | 0W4M0K0 | Creation of Vagina in Male Perineum with Nonautologous Tissue Substitute, Open Approach |
| ICD10-PCS | 0W4M0Z0 | Creation of Vagina in Male Perineum, Open Approach |
| ICD10-PCS | 0W4N071 | Creation of Penis in Female Perineum with Autologous Tissue Substitute, Open Approach |
| ICD10-PCS | 0W4N0J1 | Creation of Penis in Female Perineum with Synthetic Substitute, Open Approach |
| ICD10-PCS | 0W4N0K1 | Creation of Penis in Female Perineum with Nonautologous Tissue Substitute, Open Approach |
| ICD10-PCS | 0W4N0Z1 | Creation of Penis in Female Perineum, Open Approach |

**Table S2: National Drug Codes used to identify Emtricitabine/Tenofovir use.**

| Code type | Code | Code description |
|---|---|---|
| NDC | 61958-0701-1 | Truvada [emtricitabine/ tenofovir disoproxil fumarate], 200 mg/1, 300 mg/1 |
| NDC | 61958-0703-1 | Truvada [emtricitabine/ tenofovir disoproxil fumarate], 100 mg/1, 150 mg/1 |
| NDC | 61958-0704-1 | Truvada [emtricitabine/ tenofovir disoproxil fumarate], 133 mg/1, 200 mg/1 |
| NDC | 61958-0705-1 | Truvada [emtricitabine/ tenofovir disoproxil fumarate], 167 mg/1, 250 mg/1 |
| NDC | 61958-2005-1 | Descovy [emtricitabine and tenofovir alafenamide] 120 mg/1, 15 mg/1 |
| NDC | 61958-2002-2 | Descovy [emtricitabine and tenofovir alafenamide] 200 mg/1, 25 mg/1 |
| NDC | 61958-2002-1 | Descovy [emtricitabine and tenofovir alafenamide] 200 mg/1, 25 mg/1 |
| NDC | 70771-1620-2 | Emtricitabine / Tenofovir |
| NDC | 70771-1620-3 | Emtricitabine / Tenofovir |
| NDC | 70771-1620-4 | Emtricitabine / Tenofovir |
| NDC | 70771-1620-9 | Emtricitabine / Tenofovir |
| NDC | 70771-1621-2 | Emtricitabine / Tenofovir |
| NDC | 70771-1621-3 | Emtricitabine / Tenofovir |
| NDC | 70771-1621-4 | Emtricitabine / Tenofovir |
| NDC | 70771-1621-9 | Emtricitabine / Tenofovir |
| NDC | 70771-1622-2 | Emtricitabine / Tenofovir |
| NDC | 70771-1622-3 | Emtricitabine / Tenofovir |
| NDC | 70771-1622-4 | Emtricitabine / Tenofovir |
| NDC | 70771-1622-9 | Emtricitabine / Tenofovir |
| NDC | 70771-1709-2 | Emtricitabine / Tenofovir |
| NDC | 70771-1709-3 | Emtricitabine / Tenofovir |
| NDC | 70771-1709-4 | Emtricitabine / Tenofovir |
| NDC | 70771-1709-9 | Emtricitabine / Tenofovir |

**Table S3: Codes used to identify HIV infection.**

| Code type | Code | Code description |
|---|---|---|
| ICD10 | B20.0 | HIV disease resulting in mycobacterial infection |
| ICD10 | B20.1 | HIV disease resulting in other bacterial infections |
| ICD10 | B20.2 | HIV disease resulting in cytomegaloviral disease |
| ICD10 | B20.3 | HIV disease resulting in other viral infections |
| ICD10 | B20.4 | HIV disease resulting in candidiasis |
| ICD10 | B20.5 | HIV disease resulting in other mycoses |
| ICD10 | B20.6 | HIV disease resulting in Pneumocystis jirovecii pneumonia |
| ICD10 | B20.7 | HIV disease resulting in multiple infections |
| ICD10 | B20.8 | HIV disease resulting in other infectious and parasitic diseases |
| ICD10 | B20.9 | HIV disease resulting in unspecified infectious or parasitic disease |
| ICD10 | B21.0 | HIV disease resulting in Kaposi sarcoma |
| ICD10 | B21.1 | HIV disease resulting in Burkitt lymphoma |
| ICD10 | B21.2 | HIV disease resulting in other types of non-Hodgkin lymphoma |
| ICD10 | B21.3 | HIV disease resulting in other malignant neoplasms of lymphoid, haematopoietic and related tissue |
| ICD10 | B21.7 | HIV disease resulting in multiple malignant neoplasms |
| ICD10 | B21.8 | HIV disease resulting in other malignant neoplasms |
| ICD10 | B21.9 | HIV disease resulting in unspecified malignant neoplasm |
| ICD10 | B22.0 | HIV disease resulting in encephalopathy HIV dementia |
| ICD10 | B22.1 | HIV disease resulting in lymphoid interstitial pneumonitis |
| ICD10 | B22.2 | HIV disease resulting in wasting syndrome |
| ICD10 | B22.7 | HIV disease resulting in multiple diseases classified elsewhere |
| ICD10 | B23.0 | Acute HIV infection syndrome |
| ICD10 | B23.1 | HIV disease resulting in (persistent) generalized lymphadenopathy |
| ICD10 | B23.2 | HIV disease resulting in haematological and immunological abnormalities, not elsewhere classified |
| ICD10 | B23.8 | HIV disease resulting in other specified conditions |
| ICD10 | B24 | Unspecified human immunodeficiency virus [HIV] disease |
| ICD10 | O98.7 | HIV complicating pregnancy, childbirth, or the puerperium |
| ICD10 | O98.71X | HIV complicating pregnancy |
| ICD10 | O98.72 | HIV complicating childbirth |
| ICD10 | O98.73 | HIV complicating puerperium |
| ICD10 | Z21 | Asymptomatic HIV infection |

**Table S4: Codes used to identify HIV-associated opportunistic infections.**

| Code type | Code | Code description |
|---|---|---|
| ICD10 | A07.2 | Cryptosporidiosis |
| ICD10 | A31.0 | Pulmonary mycobacterial infection |
| ICD10 | A31.2 | Disseminated mycobacterium avium |
| ICD10 | B25.X | Cytomegaloviral disease |
| ICD10 | B37.1 | Pulmonary candidiasis |
| ICD10 | B37.81 | Candidal esophagitis |
| ICD10 | B38.X | Coccidioidomycosis |
| ICD10 | B45.X | Cryptococcosis |
| ICD10 | B58.2 | Toxoplasma meningoencephalitis |
| ICD10 | B59 | Pneumocystosis |
| ICD10 | C46.X | Kaposi's sarcoma |

### Table S5: Codes used to identify Hepatitis B infection.

| Code type | Code | Code description |
|---|---|---|
| ICD10 | B16x | Acute Hep B |
| ICD10 | B16.0 | Acute hepatitis B with delta-agent with hepatic coma |
| ICD10 | B16.1 | Acute hepatitis B with delta-agent without hepatic coma |
| ICD10 | B16.2 | Acute hepatitis B without delta-agent with hepatic coma |
| ICD10 | B16.9 | Acute hepatitis B without delta-agent and without hepatic coma |
| ICD10 | B17.0 | Acute hepatitis delta infection |
| ICD10 | B17.8 | Other specified acute viral hepatitis |
| ICD10 | B18.0 | Chronic hepatitis B with delta agent |
| ICD10 | B18.1 | Chronic hepatitis B no delta agent |
| ICD10 | B19.1 | Unspecified viral hepatitis B |
| ICD10 | B19.10 | Hepatitis B without hepatic coma |
| ICD10 | B19.11 | Hepatitis B with hepatic coma |
| ICD10 | O98.419 | Viral hepatitis complicating pregnancy, unspecified trimester |
| ICD10 | Z20.5 | Contact with and (suspected) exposure to viral hepatitis |
| ICD10 | Z11.59 | Encounter for screening for other viral diseases |
| ICD10 | Z22.51 | Hepatitis B carrier |

**Table S6: Codes used to identify needlestick exposure.**

| Code type | Code | Code description |
|---|---|---|
| ICD10 | W46 | Contact with hypodermic needle |
| ICD10 | W46.0 | Contact with hypodermic needle |
| ICD10 | W46.1 | Contact with contaminated hypodermic needle |
| ICD10 | W46.0XXA | Contact with hypodermic needle, initial encounter |
| ICD10 | W46.0XXD | Contact with hypodermic needle, subsequent encounter |
| ICD10 | W46.0XXS | Contact with hypodermic needle, sequela |
| ICD10 | W46.1XXA | Contact with contaminated hypodermic needle, initial encounter |
| ICD10 | W46.1XXD | Contact with contaminated hypodermic needle, subsequent encounter |
| ICD10 | W46.1XXS | Contact with contaminated hypodermic needle, sequela |
| ICD10 | Z29.9 | Unspecified prophylaxis |

**Table S7: Codes used to identify gonococcal infection.**

| Code type | Code | Code description |
|---|---|---|
| ICD10 | A54 | Gonococcal infection |
| ICD10 | A54.0 | Gonococcal infection of lower genitourinary tract without periurethral or accessory gland abscess |
| ICD10 | A54.00 | Gonococcal infection of lower genitourinary tract |
| ICD10 | A54.01 | Gonococcal cystitis and urethritis |
| ICD10 | A54.02 | Gonococcal vulvovaginitis |
| ICD10 | A54.03 | Gonococcal cervicitis |
| ICD10 | A54.09 | Other gonococcal infection of lower genitourinary tract |
| ICD10 | A54.1 | Gonococcal infection of lower genitourinary tract with periurethral and accessory gland abscess |
| ICD10 | A54.2 | Gonococcal pelviperitonitis and other gonococcal genitourinary infection |
| ICD10 | A54.21 | Gonococcal infection of kidney and ureter |
| ICD10 | A54.22 | Gonococcal prostatitis |
| ICD10 | A54.23 | Gonococcal infection of other male genital organs |
| ICD10 | A54.24 | Gonococcal female pelvic inflammatory disease |
| ICD10 | A54.29 | Other gonococcal genitourinary infections |
| ICD10 | A54.4 | Gonococcal infection of musculoskeletal system |
| ICD10 | A54.40 | Gonococcal infection of musculoskeletal system, unspecified |
| ICD10 | A54.41 | Gonococcal spondylopathy |
| ICD10 | A54.42 | Gonococcal arthritis |
| ICD10 | A54.43 | Gonococcal osteomyelitis |
| ICD10 | A54.49 | Gonococcal infection of other musculoskeletal tissue |
| ICD10 | A54.5 | Gonococcal pharyngitis |
| ICD10 | A54.6 | Gonococcal infection of anus and rectum |
| ICD10 | A54.8 | Other gonococcal infections |
| ICD10 | A54.81 | Gonococcal meningitis |
| ICD10 | A54.82 | Gonococcal brain abscess |
| ICD10 | A54.83 | Gonococcal heart infection |
| ICD10 | A54.84 | Gonococcal pneumonia |
| ICD10 | A54.85 | Gonococcal peritonitis |
| ICD10 | A54.86 | Gonococcal sepsis |
| ICD10 | A54.89 | Other gonococcal infections |
| ICD10 | A54.9 | Gonococcal infection |
| ICD10 | O98.211 | Gonorrhea complicating pregnancy, first trimester |
| ICD10 | O98.212 | Gonorrhea complicating pregnancy, second trimester |
| ICD10 | O98.213 | Gonorrhea complicating pregnancy, third trimester |
| ICD10 | O98.219 | Gonorrhea complicating pregnancy, unspecified trimester |
| ICD10 | O98.22 | Gonorrhea complicating childbirth |
| ICD10 | O98.23 | Gonorrhea complicating the puerperium |

**Table S8: Codes used to identify chlamydia infection.**

| Code type | Code | Code description |
|---|---|---|
| ICD10 | A55 | Chlamydial lymphogranuloma |
| ICD10 | A56.00 | Chlamydial infection of lower genitourinary tract |
| ICD10 | A56.01 | Chlamydial cystitis and urethritis |
| ICD10 | A56.02 | Chlamydial vulvovaginitis |
| ICD10 | A56.09 | Other chlamydial infection of lower genitourinary tract |
| ICD10 | A56.11 | Chlamydial female pelvic inflammatory disease |
| ICD10 | A56.19 | Other chlamydial genitourinary infection |
| ICD10 | A56.2 | Chlamydial infection of genitourinary tract, unspecified |
| ICD10 | A56.3 | Chlamydial infection of anus and rectum |
| ICD10 | A56.4 | Chlamydial infection of pharynx |
| ICD10 | A56.8 | Sexually transmitted chlamydial infection of other sites |
| ICD10 | A74.8 | Other chlamydial diseases |
| ICD10 | A74.81 | Chlamydial peritonitis |
| ICD10 | A74.9 | Chlamydial infection, unspecified |

### Table S9: Codes used to identify syphilis infection.

| Code type | Code | Code description |
|---|---|---|
| ICD10 | A51 | Early syphilis |
| ICD10 | A51.0 | Primary genital syphilis |
| ICD10 | A51.1 | Primary anal syphilis |
| ICD10 | A51.2 | Primary syphilis of other sites |
| ICD10 | A51.3 | Secondary syphilis of skin and mucous membranes |
| ICD10 | A51.31 | Condyloma latum |
| ICD10 | A51.32 | Syphilitic alopecia |
| ICD10 | A51.39 | Other secondary syphilis of skin |
| ICD10 | A51.4 | Other secondary syphilis |
| ICD10 | A51.41 | Secondary syphilitic meningitis |
| ICD10 | A51.42 | Secondary syphilitic female pelvic disease |
| ICD10 | A51.43 | Secondary syphilitic oculopathy |
| ICD10 | A51.44 | Secondary syphilitic nephritis |
| ICD10 | A51.45 | Secondary syphilitic hepatitis |
| ICD10 | A51.46 | Secondary syphilitic osteopathy |
| ICD10 | A51.49 | Gonococcal infection of eye, unspecified |
| ICD10 | A51.5 | Early syphilis latent |
| ICD10 | A51.9 | Early syphilis unspecified |
| ICD10 | A52 | Late syphilis |
| ICD10 | A52.0 | Cardiovascular and cerebrovascular syphilis |
| ICD10 | A52.00 | Cardiovascular syphilis unspecified |
| ICD10 | A52.01 | Gonococcal infection of musculoskeletal system, unspecified |
| ICD10 | A52.02 | Syphilitic aortitis |
| ICD10 | A52.03 | Syphilitic endocarditis |
| ICD10 | A52.04 | Syphilitic cerebral arteritis |
| ICD10 | A52.05 | Other cerebrovascular syphilis |
| ICD10 | A52.06 | Other syphilitic heart involvement |
| ICD10 | A52.09 | Other cardiovascular syphilis |
| ICD10 | A52.1 | Symptomatic neurosyphilis |
| ICD10 | A52.10 | Symptomatic neurosyphilis, unspecified |
| ICD10 | A52.11 | Tabes dorsalis |
| ICD10 | A52.12 | Other cerebrospinal syphilis |
| ICD10 | A52.13 | Late syphilitic meningitis |
| ICD10 | A52.14 | Late syphilitic encephalitis |
| ICD10 | A52.15 | Late syphilitic neuropathy |
| ICD10 | A52.17 | General paresis |
| ICD10 | A52.19 | Other symptomatic neurosyphilis |
| ICD10 | A52.2 | Asymptomatic neurosyphilis |
| ICD10 | A52.3 | Neurosyphilis, unspecified |
| ICD10 | A52.7 | Other symptomatic late syphilis |
| ICD10 | A52.71 | Late syphilitic oculopathy |
| ICD10 | A52.72 | Syphilis of lung and bronchus |
| ICD10 | A52.73 | Symptomatic late syphilis of other respiratory organs |
| ICD10 | A52.74 | Syphilis of liver and other viscera |
| ICD10 | A52.75 | Syphilis of kidney and ureter |
| ICD10 | A52.76 | Other genitourinary symptomatic late syphilis |
| ICD10 | A52.77 | Syphilis of bone and joint |
| ICD10 | A52.78 | Syphilis of other musculoskeletal tissue |
| ICD10 | A52.79 | Other symptomatic late syphilis |
| ICD10 | A52.8 | Late syphilis latent |
| ICD10 | A52.9 | Late syphilis unspecified |
| ICD10 | A53 | Other and unspecified syphilis |
| ICD10 | A53.0 | Latent syphilis, unspecified as early or late |
| ICD10 | A53.9 | Syphilis, unspecified |
| ICD10 | O98.1 | Syphilis complicating pregnancy, childbirth and the puerperium |
| ICD10 | O98.11 | Syphilis complicating pregnancy |
| ICD10 | O98.111 | Syphilis complicating pregnancy, first trimester |
| ICD10 | O98.112 | Syphilis complicating pregnancy, second trimester |
| ICD10 | O98.113 | Syphilis complicating pregnancy, third trimester |
| ICD10 | O98.119 | Syphilis complicating pregnancy, unspecified trimester |
| ICD10 | O98.12 | Syphilis complicating childbirth |
| ICD10 | O98.13 | Syphilis complicating the puerperium |

**Table S10: Healthcare Common Procedure Coding System (HCPCS) codes used to identify injectable antibiotics related to sexually transmitted infections.**

| Code type | Code | Code description |
|---|---|---|
| HCPCS | J0456 | Injection, azithromycin, 500 mg |
| HCPCS | J0558 | Injection, penicillin g benzathine and penicillin g procaine, 100,000 units |
| HCPCS | J0561 | Injection, penicillin g benzathine, 100,000 units |
| HCPCS | J0696 | Injection, ceftriaxone sodium, per 250 mg |
| HCPCS | J0697 | Injection, sterile cefuroxime sodium, per 750 mg |
| HCPCS | J0698 | Injection, cefotaxime sodium, per gm |
| HCPCS | J0715 | Injection, ceftizoxime sodium, per 500 mg |
| HCPCS | J0744 | Injection, ciprofloxacin for intravenous infusion, 200 mg |
| HCPCS | J1580 | Injection, garamycin, gentamicin, up to 80 mg |
| HCPCS | J2510 | Injection, penicillin g procaine, aqueous, up to 600,000 units |
| HCPCS | J2540 | Injection, penicillin g potassium, up to 600,000 units |

**Table S11: Oral and injection antibiotics with first- or second-line classification used to treat sexually transmitted infections in the United States 2017 to 2019.**

| **Antibiotic Class** | **Antibiotic Name** | **Targeted condition** | | | | | |
|---|---|---|---|---|---|---|---|
| | | *Gonorrhea*[1] | | *Chlamydia*[2] | | *Syphilis* | |
| | | First-line | Second-line | First-line | Second-line | First-line | Second-line |
| Macrolide | Azithromycin | x | x | x | x | | |
| Macrolide | Azithromycin Dihydrate | x | x | x | x | | |
| Cephalosporin | Cefixime | x | x | | | | |
| Cephalosporin | Cefotaxime Sodium | x | x | | | | |
| Quinolone | Ciprofloxacin | | x | | | | |
| Quinolone | Ciprofloxacin Hydrochloride | | x | | | | |
| Quinolone | Ciprofloxacin/Ciprofloxacin Hydrochloride | | x | | | | |
| Quinolone | Levofloxacin | | x | | | | |
| Tetracycline | Doxycycline | x | x | x | x | x | x |
| Tetracycline | Doxycycline Hyclate | x | x | x | x | x | x |
| Aminoglycoside | Gentamicin Sulfate | | x | | | | |
| Aminoglycoside | Gentamicin Sulfate/Sodium Chloride | | x | | | | |
| Penicillin | Penicillin G Benzathine | | | | | x | x |
| Penicillin | Penicillin G Benzathine/Penicillin G Procaine | | | | | x | x |
| Penicillin | Penicillin G Potassium | | | | | x | x |
| Penicillin | Penicillin G Potassium/Sodium Chloride | | | | | x | x |
| Penicillin | Penicillin G Procaine | | | | | x | x |
| Penicillin | Penicillin G Sodium | | | | | x | x |

1; Antibiotics used to treat confirmed gonorrhea infection with and without known co-infection with chlamydia.
2: Antibiotics used to treat chlamydia infection without gonorrhea co-infection.

**Table S12: Estimated annual rates of new or casual unprotected anal sex partnerships among MSM according to risk characteristics.**

| Study for risk parameterization | Stratum | Number of partners, median (interquartile range), by prior-year history of sexually transmitted infections | | | |
|---|---|---|---|---|---|
| | | *Any history* | *≥1 STI diagnosis* | *≥2 STI diagnoses* | *≥3 STI diagnoses* |
| Jin et al., 2007[8] (primary analysis) | | | | | |
| | All MSM | 2 (1-5) | 4 (1-15) | 12 (3-27) | 22 (10-39) |
| | MSM using PrEP | 6 (2-17) | 13 (4-29) | 22 (10-38) | 27 (15-48) |
| | MSM with HIV | 3 (1-8) | 6 (2-23) | 18 (5-42) | 29 (13-59) |
| Grov et al., 2016[7] | | | | | |
| | All MSM | 2 (1-5) | 2 (1-6) | 2 (1-10) | 3 (1-22) |
| | MSM using PrEP | 6 (2-17) | 7 (3-22) | 11 (3-34) | 21 (5-63) |
| | MSM with HIV | 3 (1-8) | 3 (1-11) | 4 (1-23) | 12 (2-75) |
| Janulis et al., 2023[9] | | | | | |
| | All MSM | 2 (1-5) | 11 (3-29) | 37 (19-65) | 62 (38-97) |
| | MSM using PrEP | 6 (2-17) | 22 (9-44) | 44 (25-70) | 64 (41-95) |
| | MSM with HIV | 3 (1-8) | 18 (5-46) | 52 (28-88) | 80 (50-120) |

STI: Sexually-transmitted infection (gonorrhea, chlamydia, syphilis).
Citations for included studies (7-9) are listed in the supplemental references.

### Table S13: Demographic characteristics of the study population, by risk stratum.

| **Characteristics** | | **Proportion (%) of study population** | | | | | | | | | | | | | | |
|---|---|---|---|---|---|---|---|---|---|---|---|---|---|---|---|---|
| | | *PLWH* | | | *PrEP recipient* | | | *STI history* | | | | | | | | |
| | | | | | | | | ≥1 STI | | | ≥2 STIs | | | ≥3 STIs | | |
| *Year* | | *2017* | *2018* | *2019* | *2017* | *2018* | *2019* | *2017* | *2018* | *2019* | *2017* | *2018* | *2019* | *2017* | *2018* | *2019* |
| Total, *N* | | 8095 | 10122 | 11011 | 10679 | 12480 | 15384 | 5621 | 6688 | 7609 | 825 | 1041 | 1249 | 104 | 175 | 212 |
| Sex[1] | | | | | | | | | | | | | | | | |
| | Female | 0.1 | 0.1 | 0.2 | 0.1 | 0.2 | 0.3 | 0.1 | 0.1 | 0.3 | 0.1 | 0.1 | 0.2 | 0 | 0 | 0 |
| | Male | 99.9 | 99.9 | 99.8 | 99.9 | 99.8 | 99.7 | 99.9 | 99.9 | 99.7 | 99.9 | 99.9 | 99.8 | 100 | 100 | 100 |
| Transgender[2] | | | | | | | | | | | | | | | | |
| | Yes | 0.2 | 0.3 | 0.5 | 0.3 | 0.6 | 0.7 | 0.3 | 0.3 | 0.4 | 0.2 | 0.4 | 0.5 | 0 | 0.6 | 0.5 |
| | No | 99.8 | 99.7 | 99.5 | 99.7 | 99.4 | 99.3 | 99.7 | 99.7 | 99.6 | 99.8 | 99.6 | 99.5 | 100 | 99.4 | 99.5 |
| STIs in calendar year | | | | | | | | | | | | | | | | |
| | 0 | 93.5 | 93.3 | 93.4 | 92.9 | 91.5 | 91.4 | 0 | 0 | 0 | 0 | 0 | 0 | 0 | 0 | 0 |
| | 1 | 5.2 | 5.5 | 5.4 | 5.6 | 6.8 | 6.6 | 85.3 | 84.4 | 83.6 | 0 | 0 | 0 | 0 | 0 | 0 |
| | 2 | 0.9 | 0.9 | 0.9 | 1.2 | 1.4 | 1.5 | 12.8 | 12.9 | 13.6 | 87.4 | 83.2 | 83 | 0 | 0 | 0 |
| | 3 | 0.2 | 0.2 | 0.2 | 0.2 | 0.3 | 0.3 | 1.3 | 1.9 | 1.9 | 8.6 | 12.1 | 11.4 | 68.3 | 72 | 67 |
| | >4 | 0.1 | 0.1 | 0.1 | 0.1 | 0.1 | 0.1 | 0.6 | 0.7 | 0.9 | 4 | 4.7 | 5.6 | 31.7 | 28 | 33 |
| Age group | | | | | | | | | | | | | | | | |
| | 18-24 years | 4.5 | 4.1 | 3.8 | 7.5 | 9.1 | 9.7 | 35.1 | 33.9 | 31.6 | 30.3 | 31.2 | 28.3 | 22.1 | 22.3 | 24.1 |
| | 25-34 years | 12.5 | 13.8 | 13.9 | 22 | 26.4 | 28.8 | 25.4 | 26.6 | 28.3 | 29 | 28 | 29.1 | 26.9 | 32.6 | 28.3 |
| | 35-44 years | 18.2 | 18.7 | 18.1 | 23.7 | 24.4 | 25 | 18 | 18.1 | 19.3 | 18.8 | 21.7 | 21.9 | 20.2 | 21.7 | 22.6 |
| | 45-65 years | 64.7 | 63.4 | 64.2 | 46.8 | 40.1 | 36.6 | 21.5 | 21.5 | 20.8 | 21.9 | 19 | 20.8 | 30.8 | 23.4 | 25 |
| Region | | | | | | | | | | | | | | | | |
| | West South Central | 11.3 | 8.4 | 9.5 | 10.6 | 9.3 | 10.2 | 12.5 | 9.3 | 9.6 | 10.5 | 7.2 | 8.5 | 12.5 | 8.6 | 5.7 |
| | Mountain | 4.1 | 4.1 | 4.4 | 5.4 | 5.3 | 5.6 | 4.2 | 4.5 | 4.2 | 4.5 | 3.8 | 4 | 4.8 | 3.4 | 4.7 |
| | East South Central | 6.8 | 4.4 | 4.1 | 3.5 | 2.1 | 2.6 | 5.1 | 4.3 | 4.2 | 3.6 | 4 | 3.4 | 2.9 | 2.3 | 2.4 |
| | Middle Atlantic | 16 | 12.3 | 15.8 | 17.4 | 14.3 | 20.8 | 17.4 | 13.3 | 20.6 | 20 | 13.9 | 22.7 | 22.1 | 13.1 | 25.5 |
| | South Atlantic | 33.8 | 29.9 | 30.3 | 23.3 | 20.4 | 22 | 29.1 | 24.1 | 24.3 | 27.6 | 28 | 27.7 | 23.1 | 28.6 | 26.4 |
| | West North Central | 3.5 | 3.1 | 2.7 | 3 | 3.1 | 2.7 | 3.5 | 3.7 | 3.4 | 3.4 | 3.8 | 1.9 | 1.9 | 3.4 | 0.9 |
| | East North Central | 9.2 | 10.1 | 10.1 | 9.9 | 12.1 | 11.8 | 11.4 | 12.8 | 11.1 | 9 | 10.8 | 8.8 | 3.8 | 9.1 | 7.1 |
| | Pacific | 11.7 | 11.3 | 8 | 21.7 | 21 | 15.1 | 12.9 | 13.3 | 11.2 | 16.8 | 14.5 | 13.3 | 26 | 18.9 | 16 |
| | New England | 2.9 | 2.5 | 2.4 | 4.4 | 4.2 | 3.9 | 3.4 | 3.1 | 3.4 | 4.1 | 3 | 3 | 2.9 | 2.3 | 2.4 |
| | Missing or Unknown | 0.6 | 13.9 | 12.8 | 0.9 | 8.2 | 5.4 | 0.4 | 11.7 | 8 | 0.4 | 10.9 | 6.8 | 0 | 10.3 | 9 |

STIs: Sexually-transmitted infections (gonorrhea, chlamydia, syphilis).
1: Indicates the last sex registered in medical claims data.
2: Transgender individuals are those with a diagnosis or medical procedure indicative of transgender status.

**Table S14: Incidence rate of sexually transmitted infections (STIs) among males and transgender individuals stratified by year.**

| Cohort | | Incidence rate of STIs per 100-person years (95% CI) | | |
|---|---|---|---|---|
| | | *2017* | *2018* | *2019* |
| PLWH[1] | | | | |
| | All | 7.6 (6.9, 8.4) | 8.1 (7.4, 8.8) | 8.6 (7.9, 9.3) |
| | ≥1 STI in year prior | 30.0 (25.1, 35.9) | 31.4 (26.4, 37.2) | 37.5 (32.2, 44.0) |
| | ≥2 STIs in year prior | 50.5 (36.6, 69.7) | 55.5 (41.9, 73.1) | 74.2 (56.7, 98.0) |
| | ≥3 STIs in year prior | 76.9 (47.4, 126.6) | 60.5 (40.1, 92.2) | 136.3 (91.2, 203.0) |
| PrEP use[2] | | | | |
| | All | 10.2 (9.5, 10.9) | 10.9 (10.3, 11.7) | 11.1 (10.4, 11.7) |
| | ≥1 STI in year prior | 34.6 (30.0, 39.7) | 33.5 (29.7, 38) | 37.7 (33.7, 42.3) |
| | ≥2 STIs in year prior | 49.3 (37.7, 64.4) | 59.0 (48.1, 72.6) | 64.1 (52.8, 78.0) |
| | ≥3 STIs in year prior | 51.7 (30.1, 87.0) | 78.0 (53.7, 113.8) | 97.5 (69.0, 138.2) |
| Active PrEP use[3] | | | | |
| | All | 11.7 (10.8, 12.7) | 12.2 (11.4, 13.0) | 12.4 (11.7, 13.2) |
| | ≥1 STI in year prior | 39.3 (34.3, 45.2) | 39.7 (35.5, 44.4) | 38.8 (34.7, 43.3) |
| | ≥2 STIs in year prior | 57.9 (44.9, 74.5) | 66.2 (55.2, 79.3) | 66.8 (55.7, 80.1) |
| | ≥3 STIs in year prior | 60.7 (38.7, 94.6) | 81.1 (57.4, 114.5) | 102.4 (74.6, 140.5) |
| Consistent PrEP use[4] | | | | |
| | All | 12.0 (11.1, 13.1) | 13.1 (12.1, 14.1) | 13.5 (12.6, 14.5) |
| | ≥1 STI in year prior | 42.0 (36.2, 48.8) | 43.5 (38.4, 49.3) | 43.7 (38.7, 49.4) |
| | ≥2 STIs in year prior | 57.7 (42.8, 77.6) | 68.6 (55.7, 84.4) | 76.9 (63.5, 93.0) |
| | ≥3 STIs in year prior | 69.9 (39, 124.9) | 116.7 (84.2, 161.5) | 116.6 (85.3, 159.2) |
| STI history | | | | |
| | ≥1 STI in year prior | 16.7 (15.5, 18.1) | 17.6 (16.3, 18.9) | 18.7 (17.4, 20.1) |
| | ≥2 STIs in year prior | 35.5 (30.6, 41.3) | 37.7 (33.0, 43) | 43.8 (38.7, 49.6) |
| | ≥3 STIs in year prior | 83.6 (63.6, 110.0) | 73.0 (57.7, 92.4) | 89.6 (72.2, 111.3) |

CI: Confidence interval; STIs: Sexually-transmitted infections (gonorrhea, chlamydia, syphilis).
[1]People living with HIV (PLWH).
[2]PrEP use: filled ≥1 PrEP prescription in the previous year.
[3]Active PrEP use: ≥1 PrEP prescription in the previous three months.
[4]Consistent PrEP use: ≥1 PrEP prescription in the previous three months and ≥3 PrEP fills in the previous year.

**Table S15: Incidence rate of gonorrhea diagnoses among males and transgender individuals stratified by year.**

| Cohort | | Incidence rate of gonorrhea diagnoses per 100-person years (95% CI) | | |
|---|---|---|---|---|
| | | *2017* | *2018* | *2019* |
| PLWH[1] | | | | |
| | All | 3.0 (2.6, 3.5) | 3.4 (3.0, 3.8) | 3.5 (3.1, 3.9) |
| | ≥1 STI in year prior | 15.1 (11.7, 19.5) | 15.7 (12.8, 19.4) | 18.1 (15.0, 21.9) |
| | ≥2 STIs in year prior | 28.3 (19.0, 42.0) | 28.1 (19.6, 40.5) | 74.8 (48.7, 114.9) |
| | ≥3 STIs in year prior | 38.5 (20.9, 72.5) | 33.3 (19.2, 57.3) | 136.3 (91.2, 203.0) |
| PrEP use[2] | | | | |
| | All | 5.5 (5.0, 6.0) | 6.3 (5.8, 6.8) | 5.9 (5.5, 6.4) |
| | ≥1 STI in year prior | 20.2 (17.1, 23.9) | 21.6 (18.6, 25.1) | 22.0 (19.2, 25.1) |
| | ≥2 STIs in year prior | 31.8 (24.1, 41.9) | 40.9 (32.2, 52) | 37.3 (29.8, 46.3) |
| | ≥3 STIs in year prior | 29.7 (16.6, 52.9) | 49.9 (31, 81) | 56.8 (39.9, 80.6) |
| Active PrEP use[3] | | | | |
| | All | 6.6 (6.0, 7.2) | 7.1 (6.5, 7.7) | 6.7 (6.2, 7.2) |
| | ≥1 STI in year prior | 23.6 (20.1, 27.8) | 25.5 (22.3, 29.3) | 22.4 (19.7, 25.6) |
| | ≥2 STIs in year prior | 34.2 (26.0, 45.1) | 43.4 (35.2, 53.4) | 39.2 (31.7, 48.4) |
| | ≥3 STIs in year prior | 39.4 (24.4, 63.2) | 52.7 (35.3, 78.4) | 64.4 (46.3, 89.6) |
| Consistent PrEP use[4] | | | | |
| | All | 6.7 (6.0, 7.5) | 7.7 (7, 8.4) | 7.3 (6.7, 7.9) |
| | ≥1 STI in year prior | 25.7 (21.5, 30.7) | 27.5 (23.6, 32) | 24.2 (20.8, 28.1) |
| | ≥2 STIs in year prior | 33.6 (24, 46.8) | 43.6 (34.1, 55.8) | 45.2 (36.2, 56.4) |
| | ≥3 STIs in year prior | 37 (20.4, 67.1) | 72.6 (48.8, 107.8) | 70.3 (49.7, 99.3) |
| STI history | | | | |
| | ≥1 STI in year prior | 7.6 (6.8, 8.5) | 8.8 (8, 9.7) | 9.0 (8.2, 9.8) |
| | ≥2 STIs in year prior | 16.7 (13.8, 20.3) | 19.3 (16.3, 22.8) | 22.4 (19.3, 26) |
| | ≥3 STIs in year prior | 36.5 (25.6, 52) | 33.7 (24.8, 46) | 46.6 (37, 59.4) |

CI: Confidence interval; STIs: Sexually-transmitted infections (gonorrhea, chlamydia, syphilis).
[1]People living with HIV (PLWH).
[2]PrEP use: filled ≥1 PrEP prescription in the previous year.
[3]Active PrEP use: ≥1 PrEP prescription in the previous three months.
[4]Consistent PrEP use: ≥1 PrEP prescription in the previous three months and ≥3 PrEP fills in the previous year.

**Table S16: Incidence rate of chlamydia diagnoses among males and transgender individuals stratified by year.**

| Cohort | | Incidence rate of chlamydia diagnoses per 100-person years (95% CI) | | |
|---|---|---|---|---|
| | | *2017* | *2018* | *2019* |
| PLWH[1] | | | | |
| | All | 1.6 (1.3, 1.9) | 1.9 (1.6, 2.2) | 2 (1.7, 2.3) |
| | ≥1 STI in year prior | 7.1 (5.1, 9.9) | 8.4 (6.1, 11.4) | 11.3 (8.6, 15.1) |
| | ≥2 STIs in year prior | 15.3 (9, 25.6) | 17.2 (11.5, 25.6) | 44.2 (22.6, 86.9) |
| | ≥3 STIs in year prior | 27 (11.7, 62.9) | 15.2 (6.7, 33.9) | 136.3 (91.2, 203) |
| PrEP use[2] | | | | |
| | All | 2.8 (2.5, 3.2) | 3 (2.6, 3.4) | 3.3 (3, 3.7) |
| | ≥1 STI in year prior | 10.5 (8.2, 13.4) | 7.5 (5.9, 9.7) | 11.8 (9.8, 14.3) |
| | ≥2 STIs in year prior | 16.2 (10.2, 25.8) | 14.5 (9.6, 21.9) | 21.4 (16, 28.9) |
| | ≥3 STIs in year prior | 22.3 (7.6, 65) | 24 (14.1, 41.6) | 35.1 (21.1, 59.5) |
| Active PrEP use[3] | | | | |
| | All | 3.1 (2.7, 3.6) | 3.3 (2.9, 3.7) | 3.7 (3.4, 4.1) |
| | ≥1 STI in year prior | 12.0 (9.5, 15.2) | 9.5 (7.7, 11.7) | 12.1 (10.2, 14.5) |
| | ≥2 STIs in year prior | 20.2 (13.5, 30.2) | 17.8 (12.8, 24.9) | 21.5 (16.3, 28.3) |
| | ≥3 STIs in year prior | 18.1 (7.3, 44.2) | 24.2 (14.0, 42.0) | 30.9 (19.0, 50.2) |
| Consistent PrEP use[4] | | | | |
| | All | 3.2 (2.8, 3.7) | 3.5 (3.1, 4) | 4.1 (3.6, 4.6) |
| | ≥1 STI in year prior | 13.1 (10.1, 17) | 11.9 (9.4, 15) | 15.4 (12.8, 18.5) |
| | ≥2 STIs in year prior | 20.6 (12.7, 33.5) | 22.1 (15.1, 32.2) | 26.2 (19.6, 35) |
| | ≥3 STIs in year prior | 24.6 (7.7, 77.9) | 44 (25.8, 75.1) | 43.1 (27.7, 67.1) |
| STI history | | | | |
| | ≥1 STI in year prior | 7.3 (6.5, 8.1) | 6.7 (6.0, 7.5) | 7.5 (6.8, 8.3) |
| | ≥2 STIs in year prior | 16.6 (13.5, 20.4) | 15.5 (12.9, 18.7) | 17.2 (14.5, 20.4) |
| | ≥3 STIs in year prior | 41.2 (26.7, 64.1) | 34.3 (24.8, 47.8) | 36.3 (26.6, 49.3) |

CI: Confidence interval; STIs: Sexually-transmitted infections (gonorrhea, chlamydia, syphilis).

[1]People living with HIV (PLWH).

[2]PrEP use: filled ≥1 PrEP prescription in the previous year.

[3]Active PrEP use: ≥1 PrEP prescription in the previous three months.

[4]Consistent PrEP use: ≥1 PrEP prescription in the previous three months and ≥3 PrEP fills in the previous year.

**Table S17. Incidence rate of syphilis diagnoses among males and transgender individuals stratified by year**

| Cohort | | Incidence rate of syphilis diagnoses per 100-person years (95% CI) | | |
|---|---|---|---|---|
| | | *2017* | *2018* | *2019* |
| PLWH[1] | | | | |
| | All | 3.0 (2.6, 3.4) | 2.9 (2.6, 3.3) | 3.1 (2.8, 3.5) |
| | ≥1 STI in year prior | 7.8 (5.8, 10.5) | 7.2 (5.5, 9.4) | 8.1 (6.3, 10.4) |
| | ≥2 STIs in year prior | 7.0 (3.5, 14.3) | 10.1 (6.1, 17.1) | 16.6 (7.9, 34.3) |
| | ≥3 STIs in year prior | 11.7 (4.0, 33.9) | 12.1 (4.8, 31.1) | 136.3 (91.2, 203.0) |
| PrEP use[2] | | | | |
| | All | 2.0 (1.7, 2.2) | 1.7 (1.5, 1.9) | 1.8 (1.6, 2.1) |
| | ≥1 STI in year prior | 3.9 (2.7, 5.5) | 4.4 (3.3, 5.9) | 3.9 (3.0, 5.2) |
| | ≥2 STIs in year prior | 1.3 (0.3, 5.2) | 3.6 (1.9, 7.2) | 5.5 (3.5, 8.7) |
| | ≥3 STIs in year prior | 0 (0, 0) | 4.0 (1.0, 15.6) | 5.4 (2.1, 14.1) |
| Active PrEP use[3] | | | | |
| | All | 2.0 (1.7, 2.4) | 1.8 (1.5, 2.0) | 2.0 (1.8, 2.3) |
| | ≥1 STI in year prior | 3.7 (2.5, 5.5) | 4.6 (3.5, 6.1) | 4.2 (3.3, 5.3) |
| | ≥2 STIs in year prior | 3.4 (1.5, 7.9) | 4.9 (2.9, 8.3) | 6.1 (4.1, 9.0) |
| | ≥3 STIs in year prior | 3.3 (0.8, 13.2) | 4.1 (1.2, 13.9) | 7.0 (3.6, 13.6) |
| Consistent PrEP use[4] | | | | |
| | All | 2.1 (1.8, 2.5) | 1.9 (1.6, 2.2) | 2.2 (1.9, 2.5) |
| | ≥1 STI in year prior | 3.2 (2.0, 5.2) | 4.2 (3.0, 5.9) | 4.1 (3.0, 5.6) |
| | ≥2 STIs in year prior | 3.4 (1.3, 9.0) | 2.8 (1.2, 6.7) | 5.5 (3.2, 9.3) |
| | ≥3 STIs in year prior | 8.2 (2.1, 32.4) | 0 (0, 0) | 3.2 (0.8, 12.7) |
| STI history | | | | |
| | ≥1 STI in year prior | 1.9 (1.6, 2.3) | 2.1 (1.8, 2.4) | 2.3 (1.9, 2.6) |
| | ≥2 STIs in year prior | 2.2 (1.4, 3.5) | 2.9 (2.1, 4.1) | 4.2 (3.2, 5.4) |
| | ≥3 STIs in year prior | 5.8 (2.7, 12.5) | 5.1 (2.7, 9.7) | 6.6 (4.0, 10.9) |

CI: Confidence interval; STIs: Sexually-transmitted infections (gonorrhea, chlamydia, syphilis).

[1]People living with HIV (PLWH).

[2]PrEP use: filled ≥1 PrEP prescription in the previous year.

[3]Active PrEP use: ≥1 PrEP prescription in the previous three months.

[4]Consistent PrEP use: ≥1 PrEP prescription in the previous three months and ≥3 PrEP fills in the previous year.

**Table S18: Incidence rate of oral and injection antibiotic fills related to sexually transmitted infections (STIs) among males and transgender individuals aged 18 to 24 years from 2017 to 2019 in the United States.**

| Cohort | | Incidence of antibiotic fills related to STIs per 100-person years (95% CI) | | | | | |
|---|---|---|---|---|---|---|---|
| | | *Cephalosporins* | *Macrolides* | *Penicillins* | *Tetracyclines* | *Quinolones* | *Aminoglycosides* |
| PLWH[1] | | | | | | | |
| | All | 4.92 (3.4, 7.12) | 3.32 (1.93, 5.8) | 5.91 (4.02, 8.68) | 2.82 (1.39, 5.84) | 0.12 (0.02, 0.87) | 0 (0, 0) |
| | ≥ 1 STI year prior | 5.9 (2.37, 14.65) | 5.05 (1.45, 17.73) | 4.21 (1.15, 15.29) | 5.05 (1.37, 20.26) | 0 (0, 0) | 0 (0, 0) |
| | ≥ 2 STI year prior | 6.82 (0.99, 47.02) | 10.23 (1.48, 70.53) | 3.41 (0.49, 23.51) | 0 (0, 0) | 0 (0, 0) | 0 (0, 0) |
| | ≥ 3 STI year prior | 0 (0, 0) | 0 (0, 0) | 0 (0, 0) | 0 (0, 0) | 0 (0, 0) | 0 (0, 0) |
| PrEP[2] | | | | | | | |
| | All | 5.38 (4.49, 6.45) | 4.17 (3.14, 5.56) | 2.71 (1.95, 3.75) | 1.62 (0.93, 2.82) | 0.12 (0.04, 0.39) | 0 (0, 0) |
| | ≥ 1 STI year prior | 12.32 (8.62, 17.59) | 10.67 (6.14, 18.59) | 9.85 (5.98, 16.22) | 4.5 (1.51, 13.51) | 0 (0, 0) | 0 (0, 0) |
| | ≥ 2 STI year prior | 16.61 (9.23, 29.85) | 22.12 (10.31, 50.6) | 7.37 (2.86, 18.97) | 1.83 (0.26, 12.81) | 0 (0, 0) | 0 (0, 0) |
| | ≥ 3 STI year prior | 28.44 (12.39, 65.16) | 14.13 (2.13, 93.64) | 7.07 (1.06, 46.82) | 0 (0, 0) | 0 (0, 0) | 0 (0, 0) |
| Active PrEP[3] | | | | | | | |
| | All | 6.23 (5.12, 7.57) | 5.18 (3.88, 6.97) | 2.37 (1.68, 3.33) | 1.87 (1.03, 3.4) | 0.16 (0.05, 0.51) | 0.05 (0.01, 0.39) |
| | ≥ 1 STI year prior | 15 (10.6, 21.21) | 12.57 (7.51, 21.25) | 7.73 (4.29, 13.91) | 5.3 (1.83, 15.99) | 0 (0, 0) | 0 (0, 0) |
| | ≥ 2 STI year prior | 16.87 (8.83, 32.18) | 12.62 (4.4, 37.54) | 8.42 (3.21, 22.02) | 4.19 (0.6, 29.34) | 0 (0, 0) | 0 (0, 0) |
| | ≥ 3 STI year prior | 8.25 (1.41, 47.97) | 16.49 (2.48, 109.41) | 8.24 (1.14, 59.39) | 0 (0, 0) | 0 (0, 0) | 0 (0, 0) |
| Consistent PrEP[4] | | | | | | | |
| | All | 7.56 (5.95, 9.6) | 5.83 (4, 8.6) | 2.59 (1.67, 4.02) | 1.94 (0.84, 4.47) | 0.21 (0.05, 0.86) | 0 (0, 0) |
| | ≥ 1 STI year prior | 16.18 (10.54, 24.82) | 10.77 (5.42, 21.97) | 3.07 (1.19, 7.91) | 4.59 (1.02, 21.45) | 0 (0, 0) | 0 (0, 0) |
| | ≥ 2 STI year prior | 20.21 (10.26, 39.77) | 11.51 (3.42, 44.56) | 2.87 (0.42, 19.72) | 2.87 (0.4, 20.29) | 0 (0, 0) | 0 (0, 0) |
| | ≥ 3 STI year prior | 10.71 (1.96, 58.44) | 21.39 (3.29, 139.19) | 0 (0, 0) | 0 (0, 0) | 0 (0, 0) | 0 (0, 0) |
| STI | | | | | | | |
| | ≥ 1 STI year prior | 2.57 (2.08, 3.18) | 3.25 (2.57, 4.11) | 1.29 (0.92, 1.8) | 1.25 (0.84, 1.9) | 0.04 (0.01, 0.15) | 0.18 (0.04, 0.9) |
| | ≥ 2 STI year prior | 4.6 (3.17, 6.68) | 8.12 (5.32, 12.38) | 1.22 (0.59, 2.5) | 1.62 (0.77, 3.58) | 0 (0, 0) | 0 (0, 0) |
| | ≥ 3 STI year prior | 11.35 (5.78, 22.26) | 7.55 (2.57, 23) | 2.51 (0.65, 9.71) | 0 (0, 0) | 0 (0, 0) | 0 (0, 0) |

CI: Confidence interval; STIs: Sexually-transmitted infections (gonorrhea, chlamydia, syphilis).

[1]People living with HIV (PLWH).

[2]PrEP use: filled ≥1 PrEP prescription in the previous year.

[3]Active PrEP use: ≥1 PrEP prescription in the previous three months.

[4]Consistent PrEP use: ≥1 PrEP prescription in the previous three months and ≥3 PrEP fills in the previous year.

**Table S19: Incidence rate of oral and injection antibiotic fills related to sexually transmitted infections (STIs) among males and transgender individuals aged 25 to 34 years from 2017 to 2019 in the United States.**

| Cohort | | Incidence of antibiotic fills related to STIs per 100-person years (95% CI) | | | | | |
|---|---|---|---|---|---|---|---|
| | | *Cephalosporins* | *Macrolides* | *Penicillins* | *Tetracyclines* | *Quinolones* | *Aminoglycosides* |
| PLWH[1] | | | | | | | |
| | All | 4.71 (3.94, 5.63) | 3.43 (2.68, 4.41) | 10.96 (9.48, 12.67) | 2.2 (1.47, 3.32) | 0.1 (0.04, 0.27) | 0 (0, 0) |
| | ≥ 1 STI year prior | 12.11 (9.16, 16.01) | 7.93 (4.85, 13.4) | 22.77 (17.37, 29.83) | 5.21 (2.59, 10.71) | 0.21 (0.03, 1.47) | 0 (0, 0) |
| | ≥ 2 STI year prior | 16.76 (10.44, 26.88) | 7.53 (3.58, 18.82) | 20.95 (12.96, 33.83) | 8.35 (2.69, 26.91) | 0 (0, 0) | 0 (0, 0) |
| | ≥ 3 STI year prior | 35.88 (19.52, 65.85) | 15.92 (6.47, 39.12) | 23.89 (10.32, 55.2) | 0 (0, 0) | 0 (0, 0) | 0 (0, 0) |
| PrEP[2] | | | | | | | |
| | All | 6.53 (5.97, 7.15) | 4.96 (4.37, 5.63) | 3.56 (3.07, 4.13) | 1.79 (1.37, 2.35) | 0.06 (0.03, 0.13) | 0.06 (0.02, 0.15) |
| | ≥ 1 STI year prior | 17.06 (14.26, 20.4) | 11.22 (8.65, 14.62) | 8.02 (5.83, 11.02) | 4.19 (2.41, 7.37) | 0.18 (0.05, 0.72) | 0.09 (0.01, 0.64) |
| | ≥ 2 STI year prior | 22.09 (15.95, 30.58) | 11.82 (7.18, 19.85) | 9.06 (5.06, 16.2) | 4.71 (1.42, 16.74) | 0 (0, 0) | 0 (0, 0) |
| | ≥ 3 STI year prior | 34.99 (21.38, 57.22) | 17.46 (7.87, 44.55) | 12.22 (5.04, 29.55) | 6.96 (1.48, 34.18) | 0 (0, 0) | 0 (0, 0) |
| Active PrEP[3] | | | | | | | |
| | All | 7.56 (6.89, 8.3) | 5.96 (5.22, 6.81) | 3.7 (3.16, 4.32) | 1.89 (1.41, 2.54) | 0.05 (0.02, 0.13) | 0.04 (0.01, 0.12) |
| | ≥ 1 STI year prior | 19.13 (16.07, 22.77) | 14.68 (11.43, 18.92) | 8.66 (6.41, 11.69) | 5.27 (3.1, 9.01) | 0.21 (0.05, 0.84) | 0.1 (0.01, 0.74) |
| | ≥ 2 STI year prior | 27.57 (20.58, 36.92) | 19.74 (12.69, 31.09) | 13.76 (7.81, 24.24) | 6.4 (2.03, 20.48) | 0.46 (0.06, 3.23) | 0.46 (0.06, 3.23) |
| | ≥ 3 STI year prior | 45.29 (29.19, 70.21) | 34.46 (18.26, 73.21) | 21.52 (9.52, 48.56) | 10.71 (2.12, 57.6) | 0 (0, 0) | 0 (0, 0) |
| Consistent PrEP[4] | | | | | | | |
| | All | 8.45 (7.63, 9.37) | 6.67 (5.77, 7.72) | 4.09 (3.43, 4.88) | 2.18 (1.58, 3.02) | 0.07 (0.03, 0.19) | 0.05 (0.02, 0.16) |
| | ≥ 1 STI year prior | 20.06 (16.73, 24.04) | 15.86 (12.26, 20.59) | 9.13 (6.6, 12.62) | 5.83 (3.34, 10.25) | 0.25 (0.06, 1.01) | 0.13 (0.02, 0.89) |
| | ≥ 2 STI year prior | 30.11 (22.3, 40.62) | 21.64 (13.82, 34.39) | 13.71 (7.43, 25.26) | 7.36 (2.33, 23.54) | 0.52 (0.07, 3.71) | 0.52 (0.07, 3.71) |
| | ≥ 3 STI year prior | 50.48 (32.78, 77.68) | 38.41 (20.51, 81.07) | 23.99 (10.6, 54.17) | 11.94 (2.37, 64.12) | 0 (0, 0) | 0 (0, 0) |
| STI | | | | | | | |
| | ≥ 1 STI year prior | 5.96 (5.26, 6.76) | 5.19 (4.36, 6.19) | 5.09 (4.22, 6.14) | 2.17 (1.57, 2.99) | 0.07 (0.02, 0.17) | 0.03 (0.01, 0.13) |
| | ≥ 2 STI year prior | 10.45 (8.29, 13.16) | 9.46 (7, 12.85) | 7.09 (5.18, 9.72) | 3.64 (1.93, 6.96) | 0.1 (0.01, 0.69) | 0.1 (0.01, 0.69) |
| | ≥ 3 STI year prior | 21.3 (14.66, 30.91) | 18.24 (11.3, 30.75) | 8.5 (4.37, 16.54) | 5.45 (1.76, 18.23) | 0 (0, 0) | 0 (0, 0) |

CI: Confidence interval; STIs: Sexually-transmitted infections (gonorrhea, chlamydia, syphilis).
[1]People living with HIV (PLWH).
[2]PrEP use: filled ≥1 PrEP prescription in the previous year.
[3]Active PrEP use: ≥1 PrEP prescription in the previous three months.
[4]Consistent PrEP use: ≥1 PrEP prescription in the previous three months and ≥3 PrEP fills in the previous year.

**Table S20: Incidence rate of oral and injection antibiotic fills related to sexually transmitted infections (STIs) among males and transgender individuals aged 35 to 44 years from 2017 to 2019 in the United States.**

| Cohort | | Incidence of antibiotic fills related to STIs per 100-person years (95% CI) | | | | | |
|---|---|---|---|---|---|---|---|
| | | *Cephalosporins* | *Macrolides* | *Penicillins* | *Tetracyclines* | *Quinolones* | *Aminoglycosides* |
| PLWH[1] | | | | | | | |
| | All | 2.97 (2.45, 3.6) | 2.34 (1.76, 3.14) | 8.49 (7.2, 10.01) | 1.27 (0.83, 1.95) | 0.06 (0.02, 0.18) | 0 (0, 0) |
| | ≥ 1 STI year prior | 12.07 (8.96, 16.24) | 10.22 (6.49, 16.36) | 24.96 (19.33, 32.22) | 2.86 (1.26, 7.02) | 0.2 (0.03, 1.43) | 0 (0, 0) |
| | ≥ 2 STI year prior | 24.08 (15.78, 36.7) | 21.76 (12.02, 40.6) | 35.54 (22.37, 56.41) | 2.28 (0.58, 9) | 0 (0, 0) | 0 (0, 0) |
| | ≥ 3 STI year prior | 33.21 (17.42, 63.22) | 20.75 (9.81, 43.81) | 41.47 (18.29, 93.91) | 0 (0, 0) | 0 (0, 0) | 0 (0, 0) |
| PrEP[2] | | | | | | | |
| | All | 5.21 (4.73, 5.74) | 4.26 (3.7, 4.92) | 2.56 (2.2, 2.98) | 1.64 (1.21, 2.21) | 0.08 (0.04, 0.18) | 0.02 (0.01, 0.08) |
| | ≥ 1 STI year prior | 16.19 (13.42, 19.54) | 11.28 (8.42, 15.23) | 7.98 (6.08, 10.47) | 5.58 (3.37, 9.27) | 0 (0, 0) | 0 (0, 0) |
| | ≥ 2 STI year prior | 27.87 (20.25, 38.32) | 14.68 (8.51, 25.45) | 13.67 (8.77, 21.29) | 8.08 (3.53, 18.67) | 0 (0, 0) | 0 (0, 0) |
| | ≥ 3 STI year prior | 28.12 (16.02, 49.28) | 15.29 (5.33, 45.5) | 20.41 (8.6, 48.35) | 12.72 (3.3, 57.89) | 0 (0, 0) | 0 (0, 0) |
| Active PrEP[3] | | | | | | | |
| | All | 6.18 (5.59, 6.83) | 4.97 (4.28, 5.78) | 2.7 (2.29, 3.18) | 1.94 (1.42, 2.63) | 0.09 (0.04, 0.21) | 0.03 (0.01, 0.1) |
| | ≥ 1 STI year prior | 17.46 (14.48, 21.04) | 11.81 (8.65, 16.29) | 8.66 (6.59, 11.37) | 6.03 (3.52, 10.31) | 0 (0, 0) | 0 (0, 0) |
| | ≥ 2 STI year prior | 24.02 (17.39, 33.16) | 14.28 (8.37, 24.8) | 9.14 (5.26, 15.86) | 10.27 (4.46, 24.21) | 0 (0, 0) | 0 (0, 0) |
| | ≥ 3 STI year prior | 22.02 (11.78, 41.11) | 13.18 (4.35, 39.86) | 8.79 (3.42, 22.59) | 13.17 (4.09, 52.54) | 0 (0, 0) | 0 (0, 0) |
| Consistent PrEP[4] | | | | | | | |
| | All | 6.43 (5.76, 7.17) | 5.33 (4.55, 6.26) | 2.9 (2.44, 3.45) | 2.01 (1.43, 2.82) | 0.11 (0.05, 0.26) | 0.03 (0.01, 0.13) |
| | ≥ 1 STI year prior | 18.55 (15.31, 22.47) | 12.46 (9.08, 17.28) | 9.05 (6.87, 11.92) | 5.92 (3.31, 10.61) | 0 (0, 0) | 0 (0, 0) |
| | ≥ 2 STI year prior | 25.45 (18.44, 35.09) | 15.13 (8.87, 26.26) | 9.08 (5.1, 16.13) | 9.66 (4, 24.28) | 0 (0, 0) | 0 (0, 0) |
| | ≥ 3 STI year prior | 23.04 (12.36, 42.89) | 13.79 (4.55, 41.65) | 6.89 (2.28, 20.82) | 9.18 (2.64, 36.32) | 0 (0, 0) | 0 (0, 0) |
| STI | | | | | | | |
| | ≥ 1 STI year prior | 7.57 (6.5, 8.81) | 6.22 (5, 7.75) | 8.55 (6.37, 11.48) | 3.19 (2.3, 4.42) | 0.05 (0.01, 0.21) | 0.03 (0, 0.18) |
| | ≥ 2 STI year prior | 16.49 (12.57, 21.64) | 12.95 (8.74, 19.19) | 12.22 (9.18, 16.26) | 5.59 (3.15, 9.93) | 0 (0, 0) | 0 (0, 0) |
| | ≥ 3 STI year prior | 31.78 (20.93, 48.22) | 18.51 (10, 38.4) | 22.94 (13.9, 37.82) | 14.97 (6.53, 34.54) | 0 (0, 0) | 0 (0, 0) |

CI: Confidence interval; STIs: Sexually-transmitted infections (gonorrhea, chlamydia, syphilis).
[1]People living with HIV (PLWH).
[2]PrEP use: filled ≥1 PrEP prescription in the previous year.
[3]Active PrEP use: ≥1 PrEP prescription in the previous three months.
[4]Consistent PrEP use: ≥1 PrEP prescription in the previous three months and ≥3 PrEP fills in the previous year.

**Table S21: Incidence rate of oral and injection antibiotic fills related to sexually transmitted infections (STIs) among males and transgender individuals aged 45 to 65 years from 2017 to 2019 in the United States.**

| Cohort | | Incidence of antibiotic fills related to STIs per 100-person years (95% CI) | | | | | |
|---|---|---|---|---|---|---|---|
| | | *Cephalosporins* | *Macrolides* | *Penicillins* | *Tetracyclines* | *Quinolones* | *Aminoglycosides* |
| PLWH[1] | | | | | | | |
| | All | 1.39 (1.21, 1.6) | 0.96 (0.78, 1.18) | 4.6 (4.1, 5.16) | 0.8 (0.61, 1.06) | 0.02 (0, 0.05) | 0.01 (0, 0.04) |
| | ≥ 1 STI year prior | 10 (7.83, 12.76) | 6.58 (4.68, 9.5) | 21.88 (17.37, 27.54) | 4.58 (2.55, 8.35) | 0 (0, 0) | 0 (0, 0) |
| | ≥ 2 STI year prior | 21.05 (14.05, 31.5) | 11.68 (6.29, 24.67) | 21.02 (11.39, 38.75) | 10.88 (4.77, 27.71) | 0 (0, 0) | 0 (0, 0) |
| | ≥ 3 STI year prior | 32.39 (17.43, 60.1) | 14.9 (4.96, 50.65) | 29.83 (10.81, 82.17) | 9.92 (2.56, 38.37) | 0 (0, 0) | 0 (0, 0) |
| PrEP[2] | | | | | | | |
| | All | 2.69 (2.34, 3.08) | 2.17 (1.85, 2.55) | 3.73 (3.17, 4.39) | 0.98 (0.73, 1.3) | 0.05 (0.02, 0.1) | 0.01 (0, 0.05) |
| | ≥ 1 STI year prior | 15.23 (11.2, 20.7) | 11.02 (8.27, 14.77) | 12.32 (9.15, 16.58) | 2.48 (1.25, 4.99) | 0.21 (0.05, 0.86) | 0 (0, 0) |
| | ≥ 2 STI year prior | 28.39 (15.2, 52.96) | 15.04 (8.63, 26.6) | 8.9 (4.52, 17.52) | 6.67 (2.82, 17.54) | 0.55 (0.08, 3.83) | 0 (0, 0) |
| | ≥ 3 STI year prior | 46.02 (13.22, 159.83) | 16.95 (4.8, 60.9) | 0 (0, 0) | 12.12 (3.86, 38.65) | 2.41 (0.36, 16) | 0 (0, 0) |
| Active PrEP[3] | | | | | | | |
| | All | 3.1 (2.68, 3.58) | 2.57 (2.18, 3.05) | 3.56 (2.96, 4.29) | 1.08 (0.79, 1.49) | 0.06 (0.03, 0.12) | 0.02 (0, 0.06) |
| | ≥ 1 STI year prior | 15.41 (10.79, 21.98) | 10.63 (7.61, 14.86) | 11.43 (8.57, 15.24) | 3.05 (1.48, 6.39) | 0.53 (0.2, 1.41) | 0 (0, 0) |
| | ≥ 2 STI year prior | 26.13 (12.57, 54.25) | 15.31 (8.34, 28.21) | 7.65 (4.05, 14.43) | 4.45 (1.62, 12.33) | 0.63 (0.09, 4.36) | 0 (0, 0) |
| | ≥ 3 STI year prior | 53.74 (14.69, 196.05) | 29.93 (12.69, 74.42) | 2.97 (0.42, 21.05) | 8.95 (2.01, 42.91) | 0 (0, 0) | 0 (0, 0) |
| Consistent PrEP[4] | | | | | | | |
| | All | 3.11 (2.69, 3.59) | 2.61 (2.17, 3.14) | 3.93 (3.23, 4.78) | 1.14 (0.81, 1.6) | 0.05 (0.02, 0.11) | 0.02 (0, 0.08) |
| | ≥ 1 STI year prior | 14.15 (10.22, 19.58) | 11.62 (8.31, 16.28) | 11.32 (8.34, 15.37) | 3.27 (1.56, 7.01) | 0.44 (0.14, 1.38) | 0 (0, 0) |
| | ≥ 2 STI year prior | 22.68 (10.44, 49.19) | 16.31 (8.86, 30.32) | 6.38 (3.15, 12.92) | 4.95 (1.8, 13.74) | 0.7 (0.1, 4.83) | 0 (0, 0) |
| | ≥ 3 STI year prior | 57.18 (15.67, 208.08) | 31.84 (13.54, 79.17) | 0 (0, 0) | 9.52 (2.15, 45.54) | 0 (0, 0) | 0 (0, 0) |
| STI | | | | | | | |
| | ≥ 1 STI year prior | 6.46 (5.39, 7.73) | 5.29 (4.31, 6.53) | 10.48 (8.95, 12.26) | 2.24 (1.57, 3.21) | 0.11 (0.05, 0.26) | 0 (0, 0) |
| | ≥ 2 STI year prior | 14.67 (10.21, 21.07) | 10.6 (7.03, 16.24) | 9.3 (6.5, 13.29) | 5.37 (3.1, 9.44) | 0.14 (0.02, 1.02) | 0 (0, 0) |
| | ≥ 3 STI year prior | 28.78 (14.49, 57.06) | 19.17 (8.21, 45.27) | 11.06 (4.66, 26.17) | 10.32 (4.42, 25.78) | 0.73 (0.1, 5.11) | 0 (0, 0) |

CI: Confidence interval; STIs: Sexually-transmitted infections (gonorrhea, chlamydia, syphilis).
[1]People living with HIV (PLWH).
[2]PrEP use: filled ≥1 PrEP prescription in the previous year.
[3]Active PrEP use: ≥1 PrEP prescription in the previous three months.
[4]Consistent PrEP use: ≥1 PrEP prescription in the previous three months and ≥3 PrEP fills in the previous year.

**Table S22: Incidence rate of oral and injection antibiotic fills related to gonorrhea diagnoses among males and transgender individuals from 2017 to 2019 in the United States.**

| Cohort | | Incidence rate of antibiotic fills related to gonorrhea diagnoses per 100-person years (95% CI) | | | | |
|---|---|---|---|---|---|---|
| | | *Cephalosporins* | *Macrolides* | *Tetracyclines* | *Quinolones* | *Aminoglycosides* |
| PLWH[1] | | | | | | |
| | All | 2.21 (2.01, 2.43) | 1.3 (1.15, 1.46) | 0.23 (0.17, 0.3) | 0.04 (0.02, 0.07) | 0.01 (0, 0.03) |
| | ≥1 STI in year prior | 10.8 (9.25, 12.62) | 6.41 (5.21, 7.88) | 0.93 (0.57, 1.51) | 0.1 (0.03, 0.41) | 0 (0, 0) |
| | ≥2 STIs in year prior | 19.26 (14.93, 24.83) | 10.72 (7.6, 15.13) | 1.09 (0.33, 3.61) | 0 (0, 0) | 0 (0, 0) |
| | ≥3 STIs in year prior | 31.51 (21.74, 45.65) | 14.69 (8.74, 24.68) | 0 (0, 0) | 0 (0, 0) | 0 (0, 0) |
| PrEP use[2] | | | | | | |
| | All | 4.48 (4.23, 4.76) | 2.86 (2.67, 3.05) | 0.40 (0.33, 0.47) | 0.06 (0.04, 0.1) | 0.03 (0.01, 0.05) |
| | ≥1 STI in year prior | 15.92 (14.06, 18.03) | 8.92 (7.78, 10.21) | 1.34 (0.93, 1.92) | 0.13 (0.05, 0.34) | 0.03 (0, 0.22) |
| | ≥2 STIs in year prior | 25 (19.67, 31.76) | 10.82 (8.41, 13.91) | 2.63 (1.43, 4.82) | 0.14 (0.02, 1.02) | 0 (0, 0) |
| | ≥3 STIs in year prior | 35.66 (21.75, 58.42) | 11.89 (7.19, 19.64) | 4.61 (2.03, 10.45) | 0.66 (0.09, 4.57) | 0 (0, 0) |
| Active PrEP use[3] | | | | | | |
| | All | 5.27 (4.95, 5.6) | 3.42 (3.18, 3.67) | 0.45 (0.38, 0.55) | 0.07 (0.04, 0.11) | 0.03 (0.01, 0.05) |
| | ≥1 STI in year prior | 17.29 (15.21, 19.65) | 9.64 (8.37, 11.1) | 1.69 (1.2, 2.38) | 0.22 (0.1, 0.5) | 0.04 (0.01, 0.26) |
| | ≥2 STIs in year prior | 25.33 (19.72, 32.52) | 12.58 (9.73, 16.26) | 2.68 (1.38, 5.2) | 0.33 (0.08, 1.33) | 0.17 (0.02, 1.18) |
| | ≥3 STIs in year prior | 36.46 (21.6, 61.45) | 19.7 (12.74, 30.42) | 4.37 (1.76, 10.82) | 0 (0, 0) | 0 (0, 0) |
| Consistent PrEP use[4] | | | | | | |
| | All | 5.48 (5.14, 5.86) | 3.64 (3.37, 3.94) | 0.49 (0.4, 0.6) | 0.08 (0.05, 0.13) | 0.03 (0.01, 0.06) |
| | ≥1 STI in year prior | 17.65 (15.58, 19.98) | 10.48 (9.05, 12.13) | 1.72 (1.18, 2.51) | 0.22 (0.09, 0.53) | 0.04 (0.01, 0.31) |
| | ≥2 STIs in year prior | 26.06 (20.45, 33.2) | 13.79 (10.63, 17.87) | 3.01 (1.55, 5.85) | 0.38 (0.09, 1.49) | 0.19 (0.03, 1.33) |
| | ≥3 STIs in year prior | 39.8 (23.64, 66.95) | 21.5 (13.95, 33.13) | 4.77 (1.92, 11.79) | 0 (0, 0) | 0 (0, 0) |
| STI history | | | | | | |
| | ≥1 STI in year prior | 5.46 (5.04, 5.93) | 3.43 (3.13, 3.76) | 0.59 (0.48, 0.72) | 0.07 (0.04, 0.11) | 0.06 (0.02, 0.23) |
| | ≥2 STIs in year prior | 11.32 (9.67, 13.25) | 6.9 (5.79, 8.21) | 1.35 (0.94, 1.93) | 0.06 (0.02, 0.25) | 0.03 (0, 0.23) |
| | ≥3 STIs in year prior | 24.2 (18.18, 32.18) | 11.39 (8.5, 15.25) | 2.44 (1.33, 4.45) | 0.2 (0.03, 1.42) | 0 (0, 0) |

CI: Confidence interval; STIs: Sexually-transmitted infections (gonorrhea, chlamydia, syphilis).
[1]People living with HIV (PLWH).
[2]PrEP use: filled ≥1 PrEP prescription in the previous year.
[3]Active PrEP use: ≥1 PrEP prescription in the previous three months.
[4]Consistent PrEP use: ≥1 PrEP prescription in the previous three months and ≥3 PrEP fills in the previous year.

**Table S23: Incidence rate of oral and injection antibiotic fills related to chlamydia diagnoses with and without a gonorrhea co-infection from 2017 to 2019 in the United States.**

| Cohort | | Incidence rate of antibiotic fills related to STIs per 100-person years (95% CI) | | | |
|---|---|---|---|---|---|
| | | *All chlamydia cases* | | *Chlamydia cases without gonorrhea co-infection* | |
| | | Macrolides | Tetracyclines | Macrolides | Tetracyclines |
| PLWH[1] | | | | | |
| | All | 0.48 (0.39, 0.58) | 0.29 (0.23, 0.37) | 0.3 (0.24, 0.37) | 0.24 (0.18, 0.31) |
| | ≥1 STI in year prior | 2.48 (1.69, 3.63) | 1.24 (0.82, 1.87) | 1.34 (0.86, 2.1) | 0.93 (0.57, 1.51) |
| | ≥2 STIs in year prior | 3.57 (2.02, 6.31) | 3.02 (1.68, 5.41) | 1.92 (0.92, 4) | 2.47 (1.29, 4.72) |
| | ≥3 STIs in year prior | 4.19 (1.62, 10.83) | 2.09 (0.53, 8.25) | 1.04 (0.15, 7.33) | 2.09 (0.53, 8.25) |
| PrEP use[2] | | | | | |
| | All | 0.94 (0.84, 1.05) | 0.56 (0.49, 0.65) | 0.69 (0.6, 0.78) | 0.45 (0.38, 0.53) |
| | ≥1 STI in year prior | 2.77 (2.22, 3.45) | 1.65 (1.25, 2.19) | 2.23 (1.75, 2.83) | 1.3 (0.96, 1.78) |
| | ≥2 STIs in year prior | 4.24 (2.93, 6.12) | 2.34 (1.44, 3.78) | 3.51 (2.33, 5.28) | 1.46 (0.79, 2.7) |
| | ≥3 STIs in year prior | 5.28 (2.46, 11.28) | 3.96 (1.81, 8.63) | 4.61 (2.02, 10.54) | 1.97 (0.64, 6.06) |
| Active PrEP use[3] | | | | | |
| | All | 1.11 (0.99, 1.25) | 0.66 (0.57, 0.77) | 0.83 (0.73, 0.95) | 0.54 (0.46, 0.64) |
| | ≥1 STI in year prior | 3.71 (3.03, 4.55) | 2.25 (1.72, 2.95) | 2.92 (2.32, 3.68) | 1.84 (1.35, 2.49) |
| | ≥2 STIs in year prior | 5.2 (3.68, 7.34) | 3.35 (2.04, 5.48) | 3.86 (2.58, 5.76) | 2.68 (1.51, 4.74) |
| | ≥3 STIs in year prior | 6.56 (3.47, 12.36) | 5.1 (2.46, 10.54) | 5.1 (2.47, 10.51) | 3.64 (1.53, 8.64) |
| Consistent PrEP use[4] | | | | | |
| | All | 1.11 (0.97, 1.26) | 0.69 (0.58, 0.82) | 0.82 (0.7, 0.95) | 0.55 (0.45, 0.67) |
| | ≥1 STI in year prior | 3.58 (2.87, 4.47) | 2.3 (1.71, 3.08) | 2.83 (2.21, 3.62) | 1.86 (1.33, 2.59) |
| | ≥2 STIs in year prior | 5.1 (3.52, 7.37) | 3.39 (2, 5.75) | 3.77 (2.45, 5.8) | 2.64 (1.42, 4.91) |
| | ≥3 STIs in year prior | 7.16 (3.79, 13.49) | 4.77 (2.17, 10.48) | 5.57 (2.7, 11.47) | 3.18 (1.2, 8.38) |
| STI history | | | | | |
| | ≥1 STI in year prior | 2.03 (1.81, 2.29) | 0.96 (0.83, 1.11) | 1.46 (1.28, 1.67) | 0.76 (0.64, 0.89) |
| | ≥2 STIs in year prior | 4.94 (4.07, 6) | 1.76 (1.36, 2.29) | 3.27 (2.57, 4.17) | 1.19 (0.86, 1.63) |
| | ≥3 STIs in year prior | 8.33 (5.3, 13.08) | 2.84 (1.7, 4.75) | 5.48 (2.94, 10.2) | 1.62 (0.82, 3.23) |

CI: Confidence interval; STIs: Sexually-transmitted infections (gonorrhea, chlamydia, syphilis).
[1]People living with HIV (PLWH).
[2]PrEP use: filled ≥1 PrEP prescription in the previous year.
[3]Active PrEP use: ≥1 PrEP prescription in the previous three months.
[4]Consistent PrEP use: ≥1 PrEP prescription in the previous three months and ≥3 PrEP fills in the previous year.

**Table S24: Incidence rate of oral and injection antibiotic fills related to syphilis diagnoses from 2017 to 2019 in the United States.**

| Cohort | | Incidence rate of antibiotic fills related to syphilis diagnoses per 100-person years (95% CI) | |
|---|---|---|---|
| | | *Penicillins* | *Tetracyclines* |
| PLWH[1] | | | |
| | All | 6.17 (5.7, 6.68) | 0.66 (0.57, 0.77) |
| | ≥1 STI in year prior | 21.81 (18.87, 25.21) | 2.48 (1.8, 3.42) |
| | ≥2 STIs in year prior | 23.1 (17.1, 31.19) | 3.57 (2.01, 6.33) |
| | ≥3 STIs in year prior | 29.38 (16.8, 51.32) | 2.09 (0.53, 8.25) |
| PrEP use[2] | | | |
| | All | 3.33 (3.03, 3.66) | 0.55 (0.47, 0.63) |
| | ≥1 STI in year prior | 9.42 (7.97, 11.13) | 1.46 (1.1, 1.95) |
| | ≥2 STIs in year prior | 10.23 (7.54, 13.87) | 1.9 (1.11, 3.25) |
| | ≥3 STIs in year prior | 10.56 (5.77, 19.3) | 2.63 (1, 6.89) |
| Active PrEP use[3] | | | |
| | All | 3.3 (2.98, 3.66) | 0.57 (0.49, 0.67) |
| | ≥1 STI in year prior | 9.38 (7.98, 11.02) | 1.35 (0.97, 1.87) |
| | ≥2 STIs in year prior | 10.4 (7.41, 14.58) | 1.51 (0.79, 2.88) |
| | ≥3 STIs in year prior | 11.66 (6.45, 21.06) | 2.18 (0.71, 6.7) |
| Consistent PrEP use[4] | | | |
| | All | 3.64 (3.25, 4.08) | 0.62 (0.52, 0.74) |
| | ≥1 STI in year prior | 9.42 (7.91, 11.21) | 1.46 (1.04, 2.05) |
| | ≥2 STIs in year prior | 9.62 (6.59, 14.05) | 1.51 (0.75, 3.01) |
| | ≥3 STIs in year prior | 10.34 (5.21, 20.49) | 1.58 (0.4, 6.31) |
| STI history | | | |
| | ≥1 STI in year prior | 5.96 (5.3, 6.69) | 0.78 (0.66, 0.93) |
| | ≥2 STIs in year prior | 7.31 (6.12, 8.74) | 1.44 (1.06, 1.96) |
| | ≥3 STIs in year prior | 11.58 (8.02, 16.72) | 4.06 (2.54, 6.5) |

CI: Confidence interval; STIs: Sexually-transmitted infections (gonorrhea, chlamydia, syphilis).
[1]People living with HIV (PLWH).
[2]PrEP use: filled ≥1 PrEP prescription in the previous year.
[3]Active PrEP use: ≥1 PrEP prescription in the previous three months.
[4]Consistent PrEP use: ≥1 PrEP prescription in the previous three months and ≥3 PrEP fills in the previous year.

**Table S25: Incidence rate difference of sexually transmitted infections among males and transgender individuals with 100% doxycycline postexposure prophylaxis (doxyPEP) uptake.**

| Cohort | | Incidence rate difference per 100 person-years (95% CI) | | |
|---|---|---|---|---|
| | | *Gonorrhea* | *Chlamydia* | *Syphilis* |
| PLWH[1] | | | | |
| | All | -1.5 (-2.2, -0.4) | -1.5 (-1.8, -1.0) | -2.3 (-2.6, -1.9) |
| | ≥1 STI in year prior | -7.4 (-11.1, -1.8) | -7.3 (-9.4, -4.9) | -5.9 (-7.2, -4.6) |
| | ≥2 STIs in year prior | -14.3 (-22.4, -3.6) | -14.9 (-20.4, -9.5) | -8.0 (-11.1, -5.7) |
| | ≥3 STIs in year prior | -22.3 (-37.8, -5.6) | -23.3 (-38.9, -12.8) | -10.4 (-17.5, -6.1) |
| PrEP use[2] | | | | |
| | All | -2.7 (-3.9, -0.7) | -2.5 (-2.9, -1.7) | -1.4 (-1.6, -1.1) |
| | ≥1 STI in year prior | -9.6 (-14.3, -2.4) | -8.1 (-9.8, -5.4) | -3.1 (-3.9, -2.4) |
| | ≥2 STIs in year prior | -16.7 (-25.5, -4.1) | -14.4 (-18.8, -9.5) | -3.0 (-4.5, -2.0) |
| | ≥3 STIs in year prior | -22.0 (-36.6, -5.4) | -23.1 (-34.8, -13.9) | -3.0 (-6.7, -1.4) |
| Active PrEP use[3] | | | | |
| | All | -3.1 (-4.6, -0.8) | -2.9 (-3.3, -1.9) | -1.5 (-1.7, -1.2) |
| | ≥1 STI in year prior | -11.2 (-16.6, -2.7) | -10.8 (-13.1, -7.3) | -2.8 (-3.6, -2.1) |
| | ≥2 STIs in year prior | -18.1 (-27.6, -4.6) | -18.3 (-23.7, -12) | -3.3 (-5.0, -2.2) |
| | ≥3 STIs in year prior | -27.7 (-44.6, -6.8) | -30.8 (-44.7, -19) | -3.3 (-7.6, -1.5) |
| Consistent PrEP use[4] | | | | |
| | All | -3.3 (-4.8, -0.8) | -3.0 (-3.5, -2.0) | -1.6 (-1.8, -1.3) |
| | ≥1 STI in year prior | -11.5 (-17.2, -2.8) | -11.0 (-13.3, -7.4) | -3.0 (-3.8, -2.3) |
| | ≥2 STIs in year prior | -18.9 (-28.8, -4.7) | -18.9 (-24.7, -12.4) | -3.2 (-4.8, -2.0) |
| | ≥3 STIs in year prior | -28.8 (-46.6, -7.1) | -31.7 (-47.1, -19.5) | -2.4 (-6.6, -0.9) |
| STI history | | | | |
| | ≥1 STI in year prior | -3.8 (-5.7, -1.0) | -5.8 (-6.7, -3.9) | -1.6 (-1.9, -1.3) |
| | ≥2 STIs in year prior | -8.9 (-13.4, -2.2) | -13.2 (-16.0, -9.0) | -2.5 (-3.1, -1.9) |
| | ≥3 STIs in year prior | -17.9 (-27.6, -4.4) | -29.2 (-39.1, -18.8) | -4.5 (-6.6, -3.0) |

CI: Confidence interval; STIs: Sexually-transmitted infections (gonorrhea, chlamydia, syphilis).
[1]People living with HIV (PLWH).
[2]PrEP use: filled ≥1 PrEP prescription in the previous year.
[3]Active PrEP use: ≥1 PrEP prescription in the previous three months.
[4]Consistent PrEP use: ≥1 PrEP prescription in the previous three months and ≥3 PrEP fills in the previous year.

**Table S26: Estimates of incidence rate difference in standardized tetracycline fills with 100% doxycycline postexposure prophylaxis (doxyPEP) uptake utilizing alternative estimates of sexual encounter frequencies.**

| Cohort | | Incidence rate difference for tetracycline fill-days related to STIs per 100 person-years (95% CI) | |
|---|---|---|---|
| | | *Reference manuscript* | |
| | | Grov et al., 2016 | Janulis et al., 2023 |
| PLWH | | | |
| | All | 141.0 (140.9, 141.2) | 142.1 (142.0, 142.4) |
| | ≥1 STI year prior | 188.0 (187.0, 188.9) | 2401.8 (2401.2, 2403.1) |
| | ≥2 STI year prior | 319.1 (315.3, 321.6) | 2833.2 (2832.0, 2837.0) |
| | ≥3 STI year prior | 659.1 (649.5, 661.5) | 3242.6 (3241.9, 3245.6) |
| PrEP use | | | |
| | All | 203.5 (203.5, 203.7) | 204.9 (204.8, 205.2) |
| | ≥1 STI year prior | 251.1 (250.6, 251.7) | 2147.7 (2147.1, 2148.9) |
| | ≥2 STI year prior | 359.2 (357.7, 360.6) | 2452.2 (2450.9, 2455.7) |
| | ≥3 STI year prior | 578.4 (572.8, 581.4) | 2761.1 (2758.5, 2769.7) |
| STI history | | | |
| | ≥1 STI year prior | 113.7 (113.5, 113.9) | 2275.9 (2275.7, 2276.4) |
| | ≥2 STI year prior | 169.7 (169.2, 170.3) | 2714.2 (2713.6, 2715.4) |
| | ≥3 STI year prior | 316.9 (314.3, 319.0) | 2753.8 (2752.3, 2757.5) |

CI: Confidence interval; STIs: Sexually-transmitted infections (gonorrhea, chlamydia, syphilis).
Incidence rates and incidence rate differences are reported per 100 person-years. We equated a 200 mg doxycycline dose to one-seventh of standardized tetracycline fill, reflecting the standard seven-day treatment regimen for chlamydia. The incidence rate difference in standardized tetracycline fills is equal to the increase in standardized tetracycline fills for those on doxyPEP, less STI-related tetracycline fills prevented by doxyPEP. Due to limited empirical estimates among transgender individuals, estimates are derived from men who have sex with men studies[1,10–13]. See **Table S12** for the estimated annual rates of new or casual unprotected anal sex partnerships utilizing each reference manuscript. All other tables use data from the Jin et al. study to estimate doxycycline use with doxyPEP implementation.

**Table S27. Incidence rate difference of oral and injection antibiotic fills related to sexually transmitted infections among males and transgender individuals with doxycycline postexposure prophylaxis having no effect on gonorrhea infections.**

| Cohort | | Incidence rate difference of antibiotic fills related to STIs per 100 person-years (95% CI) | | | |
|---|---|---|---|---|---|
| | | *Cephalosporins* | *Macrolides* | *Penicillins* | *Tetracyclines* |
| PLWH[1] | | | | | |
| | All | 0 (0, 0) | -0.2 (-0.2, -0.2) | -4.8 (-5.5, -3.8) | 141.0 (140.9, 141.2) |
| | ≥1 STI in year prior | 0 (0, 0) | -1.1 (-1.2, -0.8) | -16.7 (-20.2, -13.1) | 271.9 (270.9, 272.8) |
| | ≥2 STIs in year prior | 0 (0, 0) | -1.6 (-2.2, -0.9) | -17.7 (-24.4, -12.4) | 461.1 (457.3, 463.6) |
| | ≥3 STIs in year prior | 0 (0, 0) | -0.8 (-4.0, -0.1) | -22.4 (-39.6, -12.5) | 647.7 (638.1, 650.1) |
| PrEP use[2] | | | | | |
| | All | 0 (0, 0) | -0.6 (-0.6, -0.4) | -2.6 (-.03, -2.1) | 203.5 (203.4, 203.7) |
| | ≥1 STI in year prior | 0 (0, 0) | -1.8 (-1.8, -1.5) | -7.2 (-8.9, -5.6) | 312.9 (312.4, 313.5) |
| | ≥2 STIs in year prior | 0 (0, 0) | -2.8 (-3.1, -2.1) | -7.8 (-10.9, -5.5) | 421.4 (419.9, 422.8) |
| | ≥3 STIs in year prior | 0 (0, 0) | -3.7 (-5.8, -1.8) | -8.1 (-14.8, -4.3) | 532.4 (526.8, 535.4) |
| Active PrEP use[3] | | | | | |
| | All | 0 (0, 0) | -0.7 (-0.7, -0.5) | -2.5 (-3.0, -2.0) | 203.4 (203.3, 203.6) |
| | ≥1 STI in year prior | 0 (0, 0) | -2.3 (-2.4, -2.0) | -7.2 (-8.8, -5.6) | 312.4 (311.8, 313.2) |
| | ≥2 STIs in year prior | 0 (0, 0) | -3.1 (-3.3, -2.4) | -7.9 (-11.4, -5.4) | 420.7 (418.5, 422.5) |
| | ≥3 STIs in year prior | 0 (0, 0) | -4.1 (-5.7, -2.3) | -8.9 (-16.2, -4.8) | 531.5 (524.8, 535.1) |
| Consistent PrEP use[4] | | | | | |
| | All | 0 (0, 0) | -0.7 (-0.7, -0.5) | -2.8 (-3.3, -2.2) | 203.4 (203.2, 203.5) |
| | ≥1 STI in year prior | 0 (0, 0) | -2.3 (-2.3, -1.9) | -7.2 (-8.9, -5.6) | 312.3 (311.6, 313.1) |
| | ≥2 STIs in year prior | 0 (0, 0) | -3.1 (-3.3, -2.3) | -7.4 (-10.9, -4.8) | 420.6 (418.2, 422.5) |
| | ≥3 STIs in year prior | 0 (0, 0) | -4.5 (-6.3, -2.5) | -7.9 (-15.7, -3.9) | 532.2 (525., 535.50) |
| STI history | | | | | |
| | ≥1 STI in year prior | 0 (0, 0) | -1.2 (-1.2, -0.9) | -4.6 (-5.4, -3.6) | 174.6 (174.5, 174.9) |
| | ≥2 STIs in year prior | 0 (0, 0) | -2.6 (-2.7, -2.2) | -5.6 (-6.9, -4.3) | 299.2 (298.7, 299.7) |
| | ≥3 STIs in year prior | 0 (0, 0) | -4.4 (-5.6, -2.7) | -8.9 (-13.0, -5.9) | 447.2 (444.6, 449.2) |

CI: Confidence interval; STIs: Sexually-transmitted infections (gonorrhea, chlamydia, syphilis).
[1]People living with HIV (PLWH).
[2]PrEP use: filled ≥1 PrEP prescription in the previous year.
[3]Active PrEP use: ≥1 PrEP prescription in the previous three months.
[4]Consistent PrEP use: ≥1 PrEP prescription in the previous three months and ≥3 PrEP fills in the previous year.

**Table S28: Estimated increase in standardized tetracycline fills per sexually transmitted infection prevented.**

| DoxyPEP eligible population | Cohort | Excess standardized tetracycline fill-days per outcome prevented (95% CI) | | |
|---|---|---|---|---|
| | | *Gonorrhea* | *Chlamydia* | *Syphilis* |
| Entire cohort | | | | |
| | PLWH | 93.9 (69.2, 292.1) | 95.7 (80.4, 141.9) | 61.0 (53.5, 75.8) |
| | PrEP use | 76.0 (54.3, 243.2) | 82.5 (71.1, 121.6) | 145.3 (126.9, 180.3) |
| | Active PrEP use | 64.8 (46.4, 206.8) | 71.2 (61.2, 105.0) | 139.9 (121.6, 173.8) |
| | Consistent PrEP use | 61.7 (44.4, 195.8) | 69.0 (58.9, 101.9) | 128.8 (111.3, 160.3) |
| ≥ 1 STI in year prior | | | | |
| | PLWH | 36.5 (28.4, 108.0) | 37.0 (29.0, 56.2) | 46.1 (37.6, 59.3) |
| | PrEP use | 32.2 (24.1, 98.8) | 38.7 (31.8, 57.9) | 100.2 (81.4, 129.3) |
| | Active PrEP use | 27.8 (20.8, 85.4) | 28.9 (23.9, 43.0) | 109.9 (87.5, 144.0) |
| | Consistent PrEP use | 26.9 (20.2, 81.9) | 28.4 (23.3, 42.4) | 103.7 (81.9, 136.8) |
| | STI History | 45.2 (32.7, 142.9) | 30.2 (26.1, 44.6) | 108.5 (93.4, 135.7) |
| ≥ 2 STIs in year prior | | | | |
| | PLWH | 31.9 (27.0, 87.1) | 30.9 (22.3, 48.7) | 57.8 (41.6, 82.3) |
| | PrEP use | 25.0 (20.1, 72.0) | 29.2 (22.3, 44.9) | 139.9 (94.7, 211.2) |
| | Active PrEP use | 22.9 (18.3, 66.3) | 23.0 (17.7, 35.2) | 126.4 (84.8, 192.4) |
| | Consistent PrEP use | 22.0 (17.7, 63.3) | 22.2 (17.0, 34.1) | 132.7 (86.2, 208.2) |
| | STI History | 33.2 (25.2, 100.5) | 22.6 (18.7, 33.6) | 121.8 (97.0, 159.8) |
| ≥ 3 STIs in year prior | | | | |
| | PLWH | 28.4 (26.2, 70.1) | 27.8 (16.6, 49.7) | 62.2 (36.8, 106.3) |
| | PrEP use | 23.7 (21.6, 60.1) | 23.0 (15.2, 38.3) | 176.1 (79.7, 392.9) |
| | Active PrEP use | 18.9 (16.7, 49.5) | 17.3 (11.8, 28.1) | 159.2 (70.9, 360.6) |
| | Consistent PrEP use | 18.2 (16.2, 47.3) | 16.8 (11.3, 27.4) | 219.0 (82.0, 593.2) |
| | STI History | 24.8 (20.2, 69.9) | 15.3 (11.4, 23.6) | 99.3 (68.1, 148.0) |

CI: Confidence interval; STIs: Sexually-transmitted infections (gonorrhea, chlamydia, syphilis).
Estimates can be interpreted as the cohort-wide increase in tetracycline fill-days per incident STI case prevented, with doxyPEP targeted to the indicated eligible population.
[1]People living with HIV (PLWH).
[2]PrEP use: filled ≥1 PrEP prescription in the previous year.
[3]Active PrEP use: ≥1 PrEP prescription in the previous three months.
[4]Consistent PrEP use: ≥1 PrEP prescription in the previous three months and ≥3 PrEP fills in the previous year.
